# Computational modelling of EEG and fMRI paradigms reveals a consistent loss of pyramidal cell synaptic gain in schizophrenia

**DOI:** 10.1101/2021.01.07.21249389

**Authors:** Rick A Adams, Dimitris Pinotsis, Konstantinos Tsirlis, Leonhardt Unruh, Aashna Mahajan, Ana Montero Horas, Laura Convertino, Ann Summerfelt, Hemalatha Sampath, Xiaoming Michael Du, Peter Kochunov, Jie Lisa Ji, Grega Repovs, John D Murray, Karl J Friston, L Elliot Hong, Alan Anticevic

## Abstract

Diminished synaptic gain – the sensitivity of postsynaptic responses to neural inputs – may be a fundamental synaptic pathology in schizophrenia. Evidence for this is indirect, however. Furthermore, it is unclear whether pyramidal cells or interneurons (or both) are affected, or how these deficits relate to symptoms. Participants with schizophrenia (Scz, n=108), their relatives (n=57), and controls (n=107) underwent three electroencephalography paradigms – resting, mismatch negativity, and 40 Hz auditory steady-state response – and resting functional magnetic resonance imaging. Dynamic causal modelling was used to quantify synaptic connectivity in cortical microcircuits. Across all four paradigms, characteristic Scz data features were best explained by models with greater self-inhibition (decreased synaptic gain), in pyramidal cells. Furthermore, disinhibition in auditory areas predicted abnormal auditory perception (and positive symptoms) in Scz, in three paradigms. Thus, psychotic symptoms of Scz may result from a downregulation of inhibitory interneurons that may compensate for diminished postsynaptic gain in pyramidal cells.

## Introduction

Reduced excitatory synaptic gain (i.e. decreased slope of the presynaptic input-postsynaptic response relationship) is believed to be a primary deficit in schizophrenia^1,2^. This reduction may primarily affect pyramidal cells^1^ or inhibitory interneurons^3^. Decreased interneuron function in the disorder may thus be primary or a compensatory response to try to rebalance excitatory and inhibitory transmission in cortical circuits^4^. These hypotheses are difficult to assess *in vivo*, however.

Various mechanisms may reduce synaptic gain in schizophrenia: the most important is probably hypofunction of *N*-methyl-D-aspartate receptors (NMDARs) and their postsynaptic signalling cascade^1,2^. Evidence for this comes from psychiatric genetics^5^, magnetic resonance spectroscopy (MRS) imaging^6^, neuropathological studies^7^, animal models^8^, etc, but of these, only MRS is performed in humans *in vivo*, and its glutamatergic measures are difficult to interpret. Other neuromodulatory dysfunctions in schizophrenia (e.g. reduced cortical dopamine^9^ or muscarinic receptors^10^) can be assessed more directly using positron emission tomography (PET)), but neither MRS nor PET assess synaptic gain directly.

An alternative way to assess synaptic gain is using electroencephalography (EEG) paradigms such as the mismatch negativity (MMN, an auditory oddball paradigm^11^) and auditory steady-state response at 40 Hz (40 Hz ASSR, a paradigm inducing neural oscillations using a click train^12^), or in the ‘resting state’, measured with EEG (rsEEG) or functional magnetic resonance imaging (rsfMRI).

These paradigms are not direct indices of synaptic gain, however. Participants with schizophrenia diagnoses (Scz) show robust reductions in 40 Hz ASSR^12^ (*d*≈0.6) and MMN^11^ (*d*≈1) responses, which may relate to diminished synaptic gain and decreased gain modulation^13^ respectively. However, structural brain lesions also diminish the MMN^14^, and inhibitory dysfunction may reduce the 40 Hz ASSR^15^. Similarly, reduced ‘functional connectivity’ in rsfMRI analyses may result from changes in synaptic gain^16^, structural connectivity, or various confounds.

Neural mass models of non-invasive data can be parameterised in terms of synaptic gain, and these parameters estimated using dynamic causal modelling (DCM)^19^, thus furnishing model-based biomarkers – not just for Scz but also other ‘synaptopathies’ such as encephalitis^17^ and dementia^18^. DCM has several advantages: it can estimate subject-specific parameters, and its models can fit evoked (e.g. MMN) and induced (e.g. 40 Hz ASSR) EEG responses and rsfMRI, and thus explain responses to different paradigms in terms of common synaptic parameters. This is important because similar data features can behave in a paradigm-specific fashion: e.g. 40 Hz power shows little correlation between evoked response and ASSR paradigms^20^. Third, it is apt for hierarchical modelling, e.g. using group-level parameters to inform single-subject fits, using Parametric Empirical Bayes (PEB)^21^.

To date, DCM studies of Scz have used modest sample sizes and single paradigms, but have found reasonably consistent results, e.g. cortical disinhibition in EEG^13,22–24^ and rsfMRI^25^ and diminished contextual gain modulation^13,23,26^. Nevertheless, foundational questions remain, including: Are well-replicated group differences between Scz and controls across paradigms all ascribable to the same model parameter(s)? How do symptoms and cognitive dysfunction in Scz relate to these parameters? If key model parameters explain group differences, can they be reliably estimated from single subjects (in different paradigms) for ‘precision psychiatry’?

Here, we address these questions using rsEEG, MMN, 40 Hz ASSR and rsfMRI paradigms, alongside symptom and cognitive measures, in Scz (n=107), first degree relatives (Rel, n=57) and controls (Con, n=108).

## Results

In what follows, we first describe conventional analyses of group differences in data features for each paradigm. We then report the best explanation for these differences in terms of DCM parameters. Figure 1 summarises the analysis (excluding results), and Table S1 describes the participants. We used the DCM canonical microcircuit model (see below) to analyse the EEG paradigms. For the MMN and 40 Hz ASSR paradigms, we analysed group differences using conventional data features (event related potentials or power spectra), from which subject-specific DCM parameters were estimated. For rsfMRI, we modelled the network generating the MMN (and 40 Hz ASSR, in part). We used PEB (see Online Methods) to analyse group and individual differences in synaptic (model) parameters, with the exception of rsEEG, where characteristic group responses were modelled. We interpret greater ‘self-inhibition’ of pyramidal cells as an effective loss of pyramidal synaptic gain. Given known pathophysiology in Scz, NMDAR hypofunction seems the most likely explanation for loss of pyramidal gain, but other explanations are possible, for example an upregulation of (e.g., parvalbumin positive) interneuron function (see Online Methods for further discussion).

**Figure 1.**
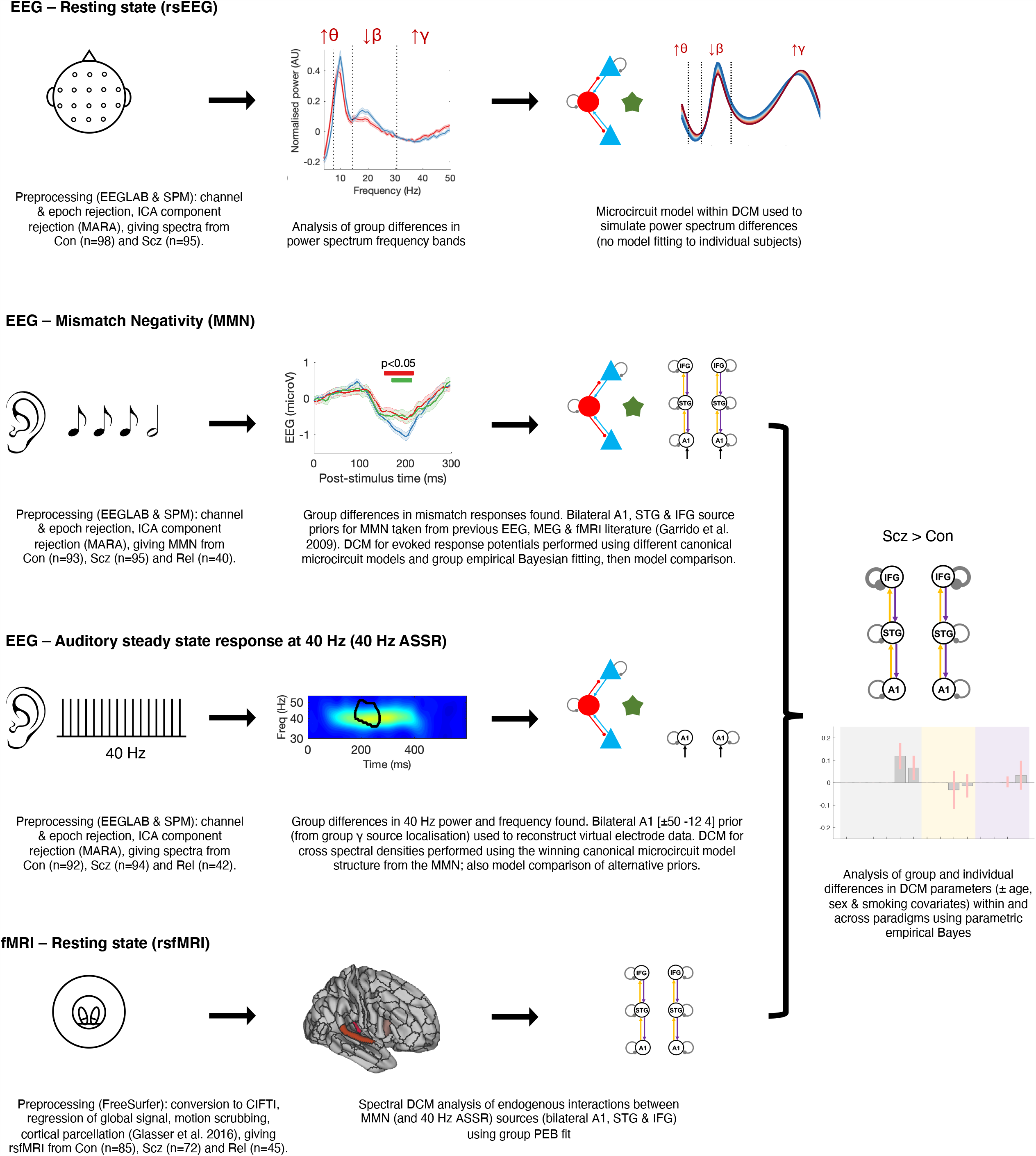
An overview of the analysis. This schematic illustrates the key steps in the preprocessing of the EEG (resting state, mismatch negativity and 40 Hz auditory steady-state response) and resting state fMRI paradigms, and their subsequent analysis using dynamic causal modelling (DCM) and parametric empirical Bayes (PEB). Simplified depictions of the paradigms are shown in the first column (see Online Methods for details), with group differences in EEG data features in the second column (first three rows), and DCM in the third column. The EEG data Con vs Scz group differences are (from first to third rows) in rsEEG θ, β, and γ frequency band power (Figure 2A), MMN responses (Figure 3A) and 40 Hz ASSR power (Figure 4C). The second column of the final row (rsfMRI) shows the Glasser parcellation areas A1 (middle), A4 (left) and 44 (right) containing the MMN sources A1, STG and IFG (respectively): these were used as nodes in the rsfMRI analysis, so that results could be compared across data modalities. Key preprocessing and analysis steps are described below the illustrations. DCM for EEG uses a cortical microcircuit model, shown on the left in the third column (also see Figure 2C). The lower three DCM illustrations include macroscopic model structures, i.e. the cortical areas involved: primary auditory cortex (A1), superior temporal gyrus (STG) and inferior frontal gyrus (IFG). In the rsEEG analysis (top row), a ‘single area’ DCM was used to reproduce power spectra characteristic of each group. In the remaining paradigms, models were fitted to the data and PEB was used to analyze group and individual differences: the final column depicts an example analysis (from Figure 3F) of group differences in DCM parameters between Con and Scz in the MMN.

Age, sex, smoking and chlorpromazine dose equivalent covariates did not significantly affect the results, unless otherwise stated. All *t*-tests were two-tailed, ranksum tests were used if distributions were skewed; none are Bonferroni-corrected unless stated.

### In rsEEG, Scz have altered power in θ, β and γ frequency bands

We first examined rsEEG power spectra by subtracting the 1/f gradient, noting that gradients did not differ between groups with eyes open or closed (*P*>0.2). The mean adjusted power spectra within the Con (n=98) and Scz (n=95) groups are shown in Figure 2A, for eyes closed (left) and open (right) conditions. The frequency bands θ (4-7 Hz), α (8-14 Hz), β (15-30 Hz) and γ (31-50 Hz) are demarcated. A repeated measures ANOVA (between-subjects factor Group, within-subjects factors Eyes open/closed and Frequency band) demonstrated a significant interaction of Frequency*Group (*F*(3, 573)=6.59, *P*<0.001) but not of Eyes*Group (*F*(1, 191)=0.05, *P*=0.8) or of Frequency*Eyes*Group (*F*(3, 573)=0.4, *P*=0.8). We therefore averaged the power in each frequency band across eyes open and closed conditions, and performed Wilcoxon ranksum tests (as the distributions were skewed), Bonferroni-corrected for four frequency bands (Figure 2B). Scz had increased θ (*Z*=2.63,

**Figure 2.**
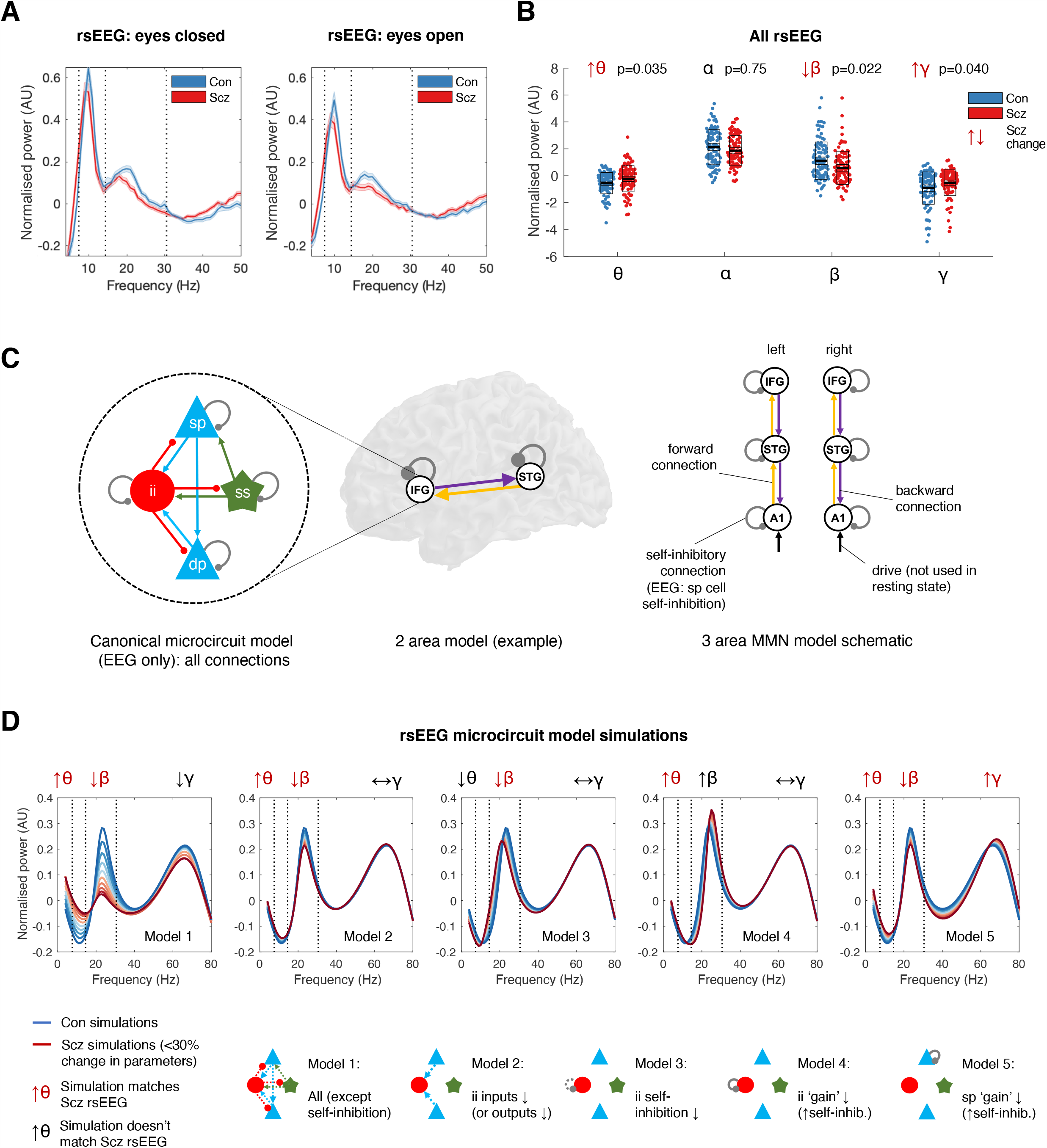
rsEEG results, DCM model structure and rsEEG simulations. A – The mean normalised eyes closed and eyes open rsEEG power spectra (±s.e.m.) across all channels for Con (n=98; blue) and Scz (n=95; red) groups, divided into four frequency bands (dotted lines): θ (3-7 Hz), α (8-14 Hz), β (15-30 Hz) and γ (>31 Hz). B – Group comparisons in mean power across both eyes closed and eyes open conditions in the θ, α, β and γ bands are shown. The box plots show the mean, s.e.m. and std. *P* values are Bonferroni-corrected for four comparisons. C – The EEG DCMs used the current version of the canonical microcircuit model^57^ (also see Figure S1A). This microcircuit (shown left) consists of superficial and deep pyramidal cells (sp and dp), inhibitory interneurons (ii), and spiny stellate (ss) cells. They are interconnected with excitatory (arrowheads) and inhibitory (beads) connections; their self-inhibitory connections parameterize their responsiveness to their inputs, i.e. synaptic gain. In EEG DCM, each modelled cortical area contains a microcircuit (middle); fMRI DCM uses a much simpler neuronal model. Both DCMs have self-inhibition parameters (round grey beads) which – in the EEG case – inhibit superficial pyramidal cells specifically. A schematic DCM diagram is explained on the right. D – The top row shows the results of five sets (Models 1-5) of simulations of microcircuit parameter changes and their similarity to the rsEEG changes in θ, β and γ bands in Scz (the model does not produce an α peak). The parameters changed in each model are illustrated in the microcircuit schematics for Models 1-5 (bottom row): parameter increases are denoted by whole lines and decreases by dotted lines. Each model is used to produce 10 simulations, starting with standard parameter values (to simulate Con) plotted in dark blue, and then reducing or increasing the parameters illustrated below in increments of 3% to simulate Scz (up to the most extreme change, plotted in dark red). Only Model 5 – a increase in superficial pyramidal self-inhibition, i.e. a loss of synaptic gain – reproduces the changes seen in all three frequency bands. *P*(corr)=0.035), decreased β (*Z*=-2.77, *P*(corr)=0.022), and increased γ (*Z*=2.58, *P*(corr)=0.040), but unchanged α (*Z*=-1.32, *P*(corr)=0.75).

### Increased pyramidal self-inhibition explains θ, β and γ changes in Scz

We used DCM’s canonical microcircuit model – a biophysical model of interacting pyramidal, interneuron and spiny stellate populations (Figure 2C, left) – to identify the most likely synaptic pathology. Model parameters include connectivity strengths (synaptic density) between populations, self-inhibition (synaptic gain) in these populations, and membrane time constants and transmission delays (Figure S1A). To model power spectrum changes in Scz, we treated cortex as a single microcircuit in which specific parameters were changed in five plausible ways (Figure 2D, bottom): a loss of all microcircuit connectivity (Model 1), a loss of pyramidal connections to or from interneurons (Model 2), interneuron disinhibition (Model 3), increased interneuron self-inhibition (Model 4) and increased pyramidal cell self-inhibition (Model 5). Note that this model does not fit the large α peak.

Only Model 5 could explain the θ, β and γ changes seen in Scz (Figure 2D, upper row); Models 1 and 2 only reproduced the θ and β changes. Model 3 showed decreased β peak frequency, which was quantitatively lower in Scz but not statistically significant (Figure S1B).

### MMN and P100 are reduced in both Scz and Rel

In the MMN paradigm, sequences of standard and duration deviant tones were presented. The ‘mismatch effect’ is the deviant–standard response in electrode Fz^11^. This difference waveform was reduced in both Scz and Rel around 200 ms (Figure 3A: Con vs Scz and Con vs Rel differences assessed using uncorrected *t* tests at each timepoint). There were no significant group differences in MMN latency between Con (mean ±std latency=194±34 ms) and Rel (196±45 ms, *P*=0.8) or Scz (202±44 ms, *P*=0.18). The mean deviant and standard waveforms are in Figure S2A: Scz showed reduced response amplitudes around 50-100 ms in both conditions, and – interestingly – an exaggerated mismatch-like response around 175 ms in the standard condition.

**Figure 3.**
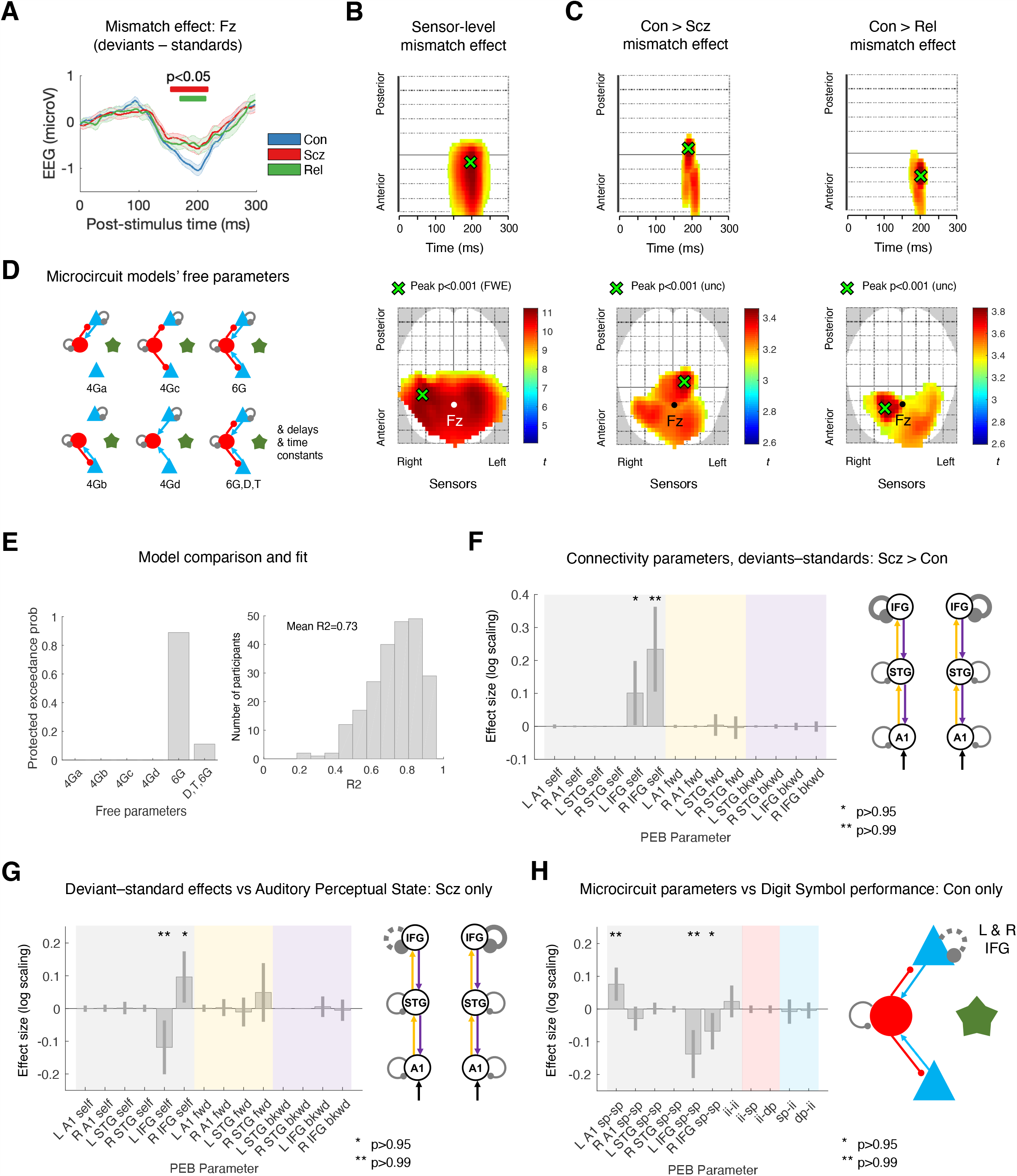
MMN data and modelling analysis. A – Mismatch difference waves (i.e. deviant–standard, mean ±s.e.m.) for Con (n=94; blue), Scz (n=96; red) and Rel (n=42; green) at electrode Fz. Group differences are computed using *t*-tests (uncorrected) at each timepoint and are marked with red (Scz vs Con) and green (Rel vs Con) bars above the difference waves. There were no significant Scz vs Rel differences. B – The lower plot shows the location of the mismatch effect (i.e. deviants – standard) at sensor level across all Con and Scz, displayed at *P*<0.05 (FWE). Fz is shown in white. The peak effect is shown in green: *P*<0.001 (FWE), *t*(376)=11.23. The upper plot shows sensors vs time: the peak effect occurs at 198 ms. C – These plots show the interaction of condition and group for the Con > Scz contrast (left) and Con > Rel contrast (right) in the same format as Fig 2B, at the lower threshold of *P*<0.005 (unc) for display purposes. Both groups demonstrate similar differences from controls in the mismatch contrast in frontocentral sensors just before 200 ms. D – Microcircuit models were compared, differing only in which parameters were allowed to change from their priors (estimated G connectivity parameters are shown, as in Figure 2C). These models’ free G parameters included various combinations of superficial (sp) and/or deep (dp) pyramidal cell (blue) connections to or from inhibitory interneurons (ii, red), and self-inhibition of sp and ii cells. Note that each parameter – within each microcircuit – could differ between subjects but was constrained to be the same in every cortical area within subjects, except for sp self-inhibition which could differ throughout. The final model also estimated delay D and time constant T parameters (these were fixed in the other five models). E – Model comparison and evaluation. Left: The protected exceedance probability is the probability a particular model is more likely than any other tested model, above and beyond chance, given the group data. The model with most free parameters is at the far right; it comes second to the 6G model with fixed delays (D) and time constants (T) and 6 microcircuit connectivity parameters estimated. Right: A histogram of R^2^ values for all participants for the winning model: it fits most participants well. F – A PEB analysis of MMN model parameters (i.e. connections) that contribute to the Scz > Con mismatch effect. The results are plotted on the left (with 95% Bayesian confidence intervals) and shown in schematic form on the right; parameters with posterior probabilities of *P*>0.95 or *P*>0.99 of contributing to the group difference effect are indicated with one or two asterisks (respectively). On the plot, self-inhibitory connections are shaded grey, forward connections shaded yellow, and backward connections shaded purple (matching the colours in the schematic). The y axis denotes log-scaling of the effect size: changes of exp(±0.2) are of roughly ±20%. Some parameters have been eliminated during Bayesian model reduction (see Online Methods). The analysis indicates Scz showed greater self-inhibition (or reduction in synaptic gain) in bilateral IFG in the mismatch contrast. The Rel > Con contrast did not show significant effects. G – A PEB analysis of MMN mismatch effect model parameters that correlate with current (‘state’) abnormal auditory percepts within Scz only, plotted in the same format as Figure 3F. Within Scz, abnormal auditory percepts relate to reduced self-inhibition in right IFG, but disinhibition in left IFG (in Broca’s area). H – A PEB analysis of MMN microcircuit model parameters that correlate with Digit Symbol task performance within Con only. The results plotted on the left correspond to microcircuit parameters (illustrated on the right): blue shaded connections are superficial and deep pyramidal inputs to interneurons, red shaded connections are interneuron outputs to pyramidal layers, and grey shaded connections are self-inhibitory. Self-inhibitory connections are estimated in interneuron and superficial pyramidal populations: the latter differ in every cortical area (all other microcircuit parameters are identical in all cortical areas). See Figure 3F’s legend for additional details. Better performance on the Digit Symbol task relates to reduced self-inhibition (i.e. increased synaptic gain) of superficial pyramidal cells in IFG (indicated using dotted lines on the schematic). N.B. All effects shown in F to H are also present without the addition of age, sex, and smoking covariates (*P*>0.95). Inclusion of a chlorpromazine dose equivalent covariate renders the analysis in 3F non-significant (*P*>0.75), but it makes the overall effect of Scz on L & R IFG self-inhibition become significant (see Figure S4C).

Smoothed sensor-level data were analysed using cluster-based statistics. Across Con and Scz, there was a strong mismatch effect, peaking at 198 ms (peak *P*_(FWE)_<0.001, *t*(376)=11.23; Figure 3B), which was reduced in Scz (peak at 186 ms, *P*_(unc)_<0.001, *t*(376)=3.46) and in Rel (peak at 198 ms, *P*_(unc)_<0.001, *t*(268)=3.83; Figure 3C). Likewise, Scz had a reduced P100 response (peak at 82 ms, *P*_(FWE)_=0.003, *t*(376)=4.83), as did Rel (peak at 94 ms, *P*_(unc)_=0.001, *t*(268)=3.02; Figure S2B).

### DCM of MMN indicates increased frontal self-inhibition in Scz, and group-specific relationships of self-inhibition with abnormal auditory percepts and cognitive performance

We first used model comparison to establish whether it was best to fix or estimate various microcircuit parameters in the MMN analysis (see Online Methods). We compared six models (Figure 3D): Model 6G estimates six connectivity (G) parameters, Models 4Ga-d consider subsets of these six, and Model 6G,D,T also estimates delays and time constants. Bayesian model selection preferred Model 6G (also in Con and Scz separately), with a protected exceedance probability of *P*=0.89 (Figure 3E, left). This model fitted most participants’ data accurately (e.g. Figure S3A): a histogram of R^2^ values is shown in Figure 3E (right) – the group mean R^2^ was 0.73. R^2^ were slightly higher in Con (mean=0.76 ±std=0.13) than in Scz (0.70±0.14; ranksum *Z*=3.12, *P*=0.0018) and Rel (0.71±0.15; ranksum *Z*=2.14, *P*=0.033) (Figure S3C).

We then used PEB to ask which parameters best explained group differences in the MMN: self-inhibition within areas or connections between areas. The reduced mismatch effect in Scz was best explained by increased self-inhibition in deviant – relative to standard – trials in L IFG (*P*>0.95) and R IFG (*P*>0.99; Figure 3F). Including chlorpromazine dose equivalent covariates reduced the posterior probability to *P*>0.75, but age, sex and smoking had no effect. Conversely, there was no overall group effect (across both standards and deviants) of Scz on the microcircuit parameters (all *P*<0.95; Figure S4C, left) unless chlorpromazine dose equivalents were included as covariates: here, Scz showed greater superficial pyramidal self-inhibition in L and R IFG (both *P*>0.99; Figure S4C, middle and right) and reduced interneuron self-inhibition throughout (*P*>0.95). Rel did not show effects of *P*>0.95 in either analysis.

In Scz, the auditory perceptual abnormalities ‘state’ measure was associated with disinhibition in L IFG (*P*>0.99) – within Broca’s area – but increased self-inhibition in R IFG (*P*>0.95) in the mismatch contrast (Figure 3G). Historical auditory perceptual abnormalities (the ‘trait’ measure) showed similar effects but at lower posterior probability (*P*>0.75, not shown). In Con, across both conditions, reduced self-inhibition in bilateral IFG was associated with greater information processing speed (i.e. Digit Symbol Substitution Task score; *P*>0.99 and *P*>0.95), as was increased self-inhibition in A1 (*P*>0.99), but there were no such relationships in Scz (all *P*<0.95).

### Scz had reduced γ power and peak frequency in 40 Hz ASSR, and Rel reduced γ power

We next considered induced responses during auditory steady-state stimulation. Group-averaged 40 Hz ASSR are shown in Figure 4A, and the distributions of participants’ peak γ (35-45 Hz) frequencies in Figure 4B. Scz had slightly reduced γ peak frequency: mean peak frequencies (following subtraction of the 1/f gradient: Figure S2D) were Con=40.2 Hz (std 1.7), Scz=39.5 Hz (std 1.7; *t*(184)=2.67, *P*=0.008) and Rel=39.9 Hz (std 2.1; *t*(132)=1.03, *P*=0.3). Adjusted time-frequency plots are shown in Figure 4C (and raw time frequency data in Figure S2E): Con showed a robust increase in ∼40 Hz power around 100 ms, which is diminished in Scz and Rel (*P*<0.05 *t*-tests at each frequency and timepoint are circled on the middle and right plots, for Con vs Scz and Con vs Rel in black and Scz vs Rel in white: this many differences are unlikely due to chance – Con vs Scz and Con vs Rel both *P*<0.001, Scz vs Rel *P*=0.006, permutation tests).

**Figure 4.**
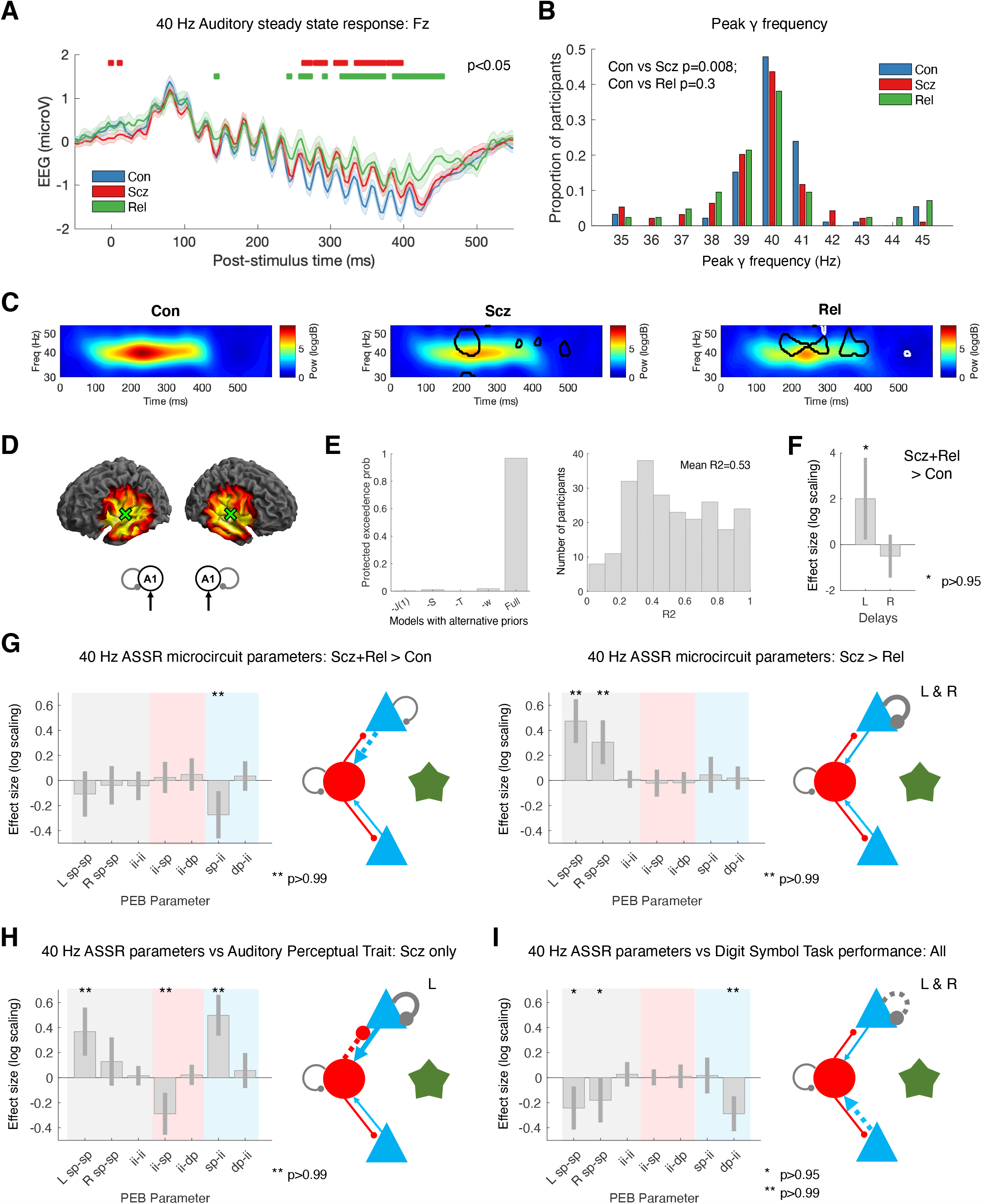
40 Hz ASSR data and modelling analysis. A – 40 Hz ASSR time courses at electrode Fz for Con (n=92; blue), Scz (n=94; red) and Rel (n=42; green). 16 clicks were played at 40 Hz, starting at 0 ms. Group differences in the baseline deflection (not modelled subsequently) emerge after around 250 ms: shown with red bars (Con vs Scz) and green bars (Con vs Rel), both *P*<0.05 (*t*-tests per timepoint, uncorrected). B – γ (35-45 Hz) frequencies with the strongest power (in the normalized spectrum) in each participant are shown in a histogram. C – These normalised time frequency plots show the ∼40 Hz responses around 100-400 ms. The Scz and Rel plots have areas of difference from Con encircled in black; the Rel plot has areas of difference from Scz encircled in white (*P*<0.05 *t*-tests at each time and frequency). D – The left plots show the bilateral A1 (transverse temporal gyrus) sources chosen following source localization: [±50 −12 4]. The 40 Hz ASSR model structure is on the right: bilateral sources in A1. E – Left: To improve the DCM fit of the cross spectral densities in bilateral A1 in this non-standard paradigm, we used empirical priors (also see Figure S1A) for: J(1), the contribution spiny stellate cells make to the EEG signal; S, the gain of the neuronal activation function; T, population time constants; and also w, the width of the ∼40 Hz Gaussian bump. The plot shows that the Full model (with all the empirical priors) is superior to other models that used standard values for their respective priors (or for ‘-w’, 1 Hz instead of 4 Hz). Right: a histogram of R^2^s for all participants for the winning model. F – PEB analysis indicated Scz+Rel > Con showed increased neural transmission delays in L A1. G – Left: PEB analysis (in the same format as Figure 3H) indicated Scz+Rel > Con (a psychosis ‘genetic risk’ effect) had decreased sp-ii connectivity. Right: Scz > Rel (a psychosis ‘diagnosis’ effect) shows decreased sp self-inhibition in bilateral A1. H – PEB analysis in Scz, showing abnormal auditory percepts are associated with disinhibition of the sp-ii circuit (and increased sp self-inhibition in L A1). I – PEB analysis across all participants, showing Digit Symbol score relates to reduced sp self-inhibition (i.e. increased synaptic gain), and lower dp-ii connectivity. All effects shown in F, G, H and I are also present without the addition of age, sex and smoking covariates (*P*>0.95), and also with inclusion of chlorpromazine dose equivalents as a covariate.

### 40 Hz ASSR DCM suggests a loss of pyramidal input to interneurons in Scz and Rel, and greater self-inhibition in Scz

The peak cortical source – closest to A1 – was [50 −12 4], hence bilateral sources at [±50 −12 4] were used as priors for reconstruction of virtual electrode data: the DCM comprised these bilateral sources and their thalamic drive (Figure 4D). Empirical priors for several parameters were used to optimise model fit (Figure S1A). Bayesian model comparison between the Full model (containing empirical priors for the contribution of spiny stellate cells to measured signals, the neural activation function, and synaptic time constants) and models with standard priors for these parameters showed the Full model was superior (Figure 4E, left). The 40 Hz thalamic drive was modelled using a Gaussian bump function of width w≤4 Hz (see Online Methods): this width performed better than a narrower bump of 1 Hz (Model -w, Figure 4E). Model fits for the winning model were reasonable (Figure S3B; mean R^2^=0.53). Group differences in R^2^ were not detected (Figure S3C, ranksum tests: all *P*>0.1).

Unlike the MMN analysis, we performed group comparisons with PEB using psychosis ‘genetic risk’ (Scz+Rel > Con) and ‘diagnosis’ (Scz > Rel) as explanatory variables^13,23^. This was because there were marked differences between Rel and Con parameters, only some of which were shared by Scz (Figure S5B). The ‘genetic risk’ effect was expressed as an increased conduction delay in L A1 (*P*>0.95; Figure 4F), and reduced superficial pyramidal (sp) to inhibitory interneuron (ii) connectivity (*P*>0.99; Figure 4G, left). The psychosis ‘diagnosis’ effect was associated with increased superficial pyramidal self-inhibition in bilateral A1 in Scz (both *P*>0.99; Figure 4G, right).

### 40 Hz ASSR DCM links abnormal auditory percepts to A1 disinhibition in Scz and cognitive performance to reduced A1 self-inhibition in all participants

In Scz, the auditory perceptual abnormalities ‘trait’ measure related to a disinhibited sp-ii-sp circuit, i.e. increased sp-ii (*P*>0.99) and reduced ii-sp connectivity (*P*>0.99), as well as greater self-inhibition in L A1 (*P*>0.99; Figure 4H). These associations were also seen in the auditory ‘state’ measure but at lower posterior probability (*P*>0.95 for sp-ii, *P*>0.75 for ii-sp and sp-sp, not shown). Across all participants, Digit Symbol score was associated with reduced pyramidal self-inhibition in bilateral A1 (both *P*>0.95) and reduced deep pyramidal to inhibitory interneuron (dp-ii) connectivity throughout (*P*>0.99; Figure 4I). Self-inhibition and dp-ii effects were also present in both Con and Scz groups when analysed separately (the former in L A1 in Con, and R A1 in Scz; both *P*>0.95, not shown).

### rsfMRI DCM of the MMN circuit finds increased self-inhibition in IFG in Scz and Rel

To generalise our inferences – based upon the DCMs of electrophysiological data – we analysed effective connectivity using haemodynamic responses within the ‘MMN network’ during rsfMRI, i.e. the Glasser parcellation areas (in the rsfMRI data) based on the MMN source locations (see Online Methods): bilateral A1, A4 and 44 (Figure 1). The microcircuit model used to explain fMRI data is simpler than the neural mass models used for EEG; however, they retain inhibitory self-connections. Model fits were accurate: R^2^s were >0.7 in all groups, with no group differences (Figure S3C, ranksum tests: all *P*>0.05).

PEB analysis of subject-specific DCM parameters suggested that Scz is associated with increased self-inhibition in L and R IFG (*P*>0.99 and *P*>0.95 respectively; Figure 5A). These effects were robust to age, sex, and smoking covariates (and to the removal of the 10 participants with the lowest rsfMRI signal to noise ratio: 8 Scz and 2 Con; both *P*>0.95). These effects did not survive addition of chlorpromazine dose equivalents (L IFG self-inhibition fell to *P*>0.75). However, Rel > Con showed the same increase in self-inhibition in bilateral IFG (both *P*>0.95, Figure 5B). This group difference did not survive addition of the age covariate: Rel were older than Con (Rel mean age=45.4 ±16.6 years, Con mean age=39.4 ±14.3 years; *t*(162)=2.4, *P*=0.02). These differences were not detected using conventional functional connectivity analyses (that cannot assess self-inhibition) or analyses of regional variance (see Figures S6B to S6E and Online Methods for further discussion).

**Figure 5.**
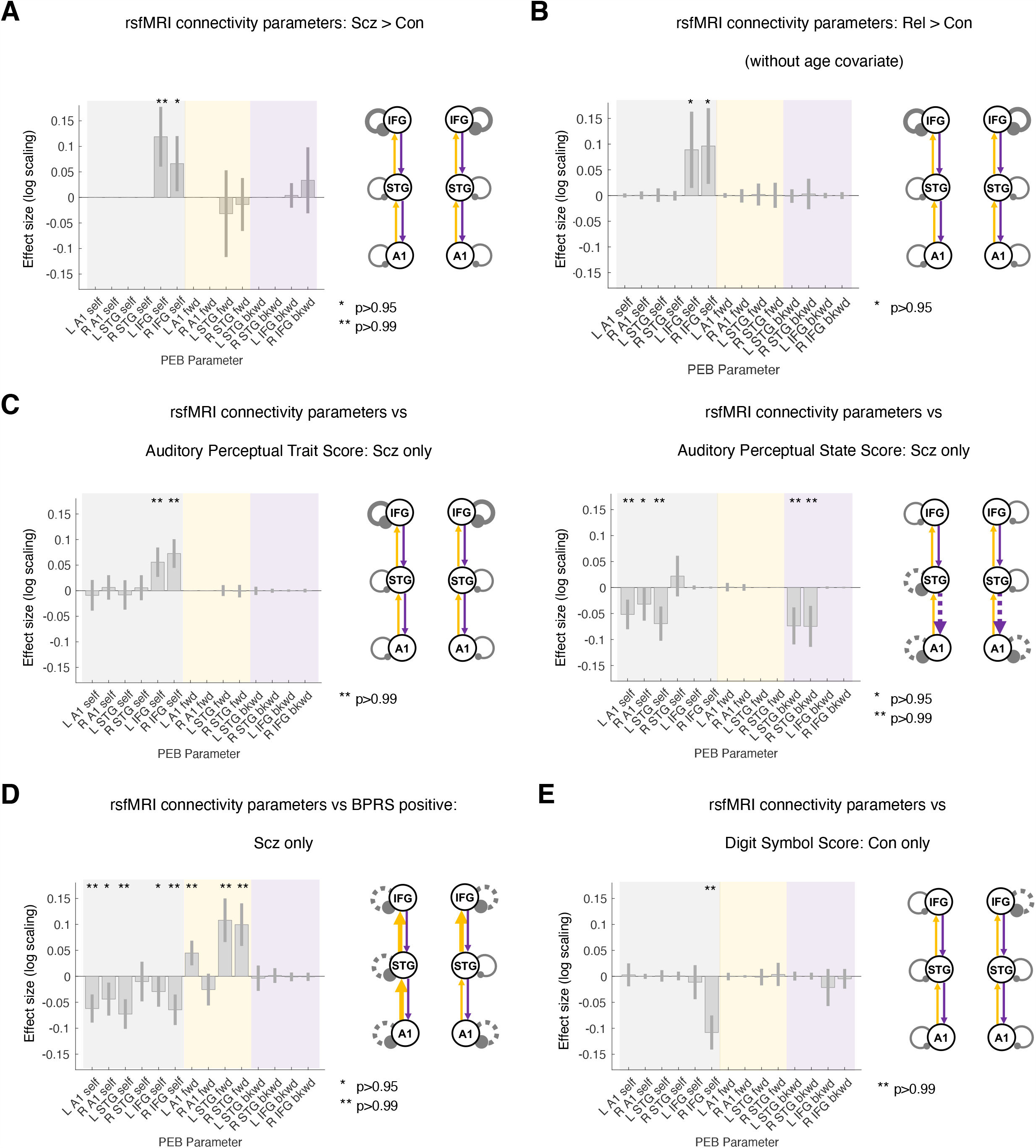
Resting state fMRI modelling analysis. A – For comparative purposes, the rsfMRI connectivity analysis was conducted on the same network as the MMN analysis. Results for Con (n=85) and Scz (n=72) are shown in the same format as Figure 3F. As in the MMN, Scz showed increased self-inhibition in bilateral IFG. Inclusion of chlorpromazine equivalent dose as a covariate still showed increased self-inhibition in L IFG but only at *P*>0.75. B – rsfMRI connectivity analysis without covariates for Con (n=85) and Rel (n=45) is shown. Like Scz, Rel show increased self-inhibition in bilateral IFG, but this effect disappeared with addition of the age covariate (*P*<0.75). C – Left: Within Scz, abnormal auditory percepts (‘trait’ measure) related to increased self-inhibition in bilateral IFG. Right: Conversely, abnormal auditory percepts (‘state’ score – i.e. experiences within the last week only) relates to disinhibition in temporal areas and also a loss of top down connections within auditory cortex. D – Within Scz, BPRS positive symptom score related to disinhibition throughout the MMN network and increased forward connectivity in 3/4 connections. E – In Con, Digit Symbol score related to reduced self-inhibition in R IFG. All effects shown (except Rel > Con) are also present without the addition of age, sex and smoking covariates, and also if participants (2 controls, 8 Scz) with rsfMRI SNR <25 are excluded (all *P*>0.95). Some rsfMRI results are no longer significant without GSR (Figure S7). No results change substantially with inclusion of chlorpromazine dose equivalent as a covariate unless stated.

### rsfMRI DCM reveals relationships of positive symptoms to cortical disinhibition in Scz and of gain to cognition in Con

PEB analysis within Scz found that the auditory perceptual abnormalities ‘trait’ measure was associated with increased self-inhibition in L and R IFG (both *P*>0.99, Figure 5C, left). Conversely, Scz auditory perceptual abnormalities ‘state’ scores were associated with disinhibition in R A1 (*P*>0.95) and L A1 and STG (both *P*>0.99), and of STG-A1 backward connectivity bilaterally (both *P*>0.99; Figure 5C, right). The R A1 effect was attenuated if age, sex, and smoking covariates were not included, and if a chlorpromazine dose equivalent covariate was added.

Similarly, BPRS positive symptoms (including age, sex, smoking and negative symptoms covariates) were associated with decreased self-inhibition everywhere except R STG (all *P*>0.99 except L IFG and R A1, both *P*>0.95) and stronger forward connections everywhere except R Al-STG (all *P*>0.99; Figure 5D). Most effects were robust to addition of chlorpromazine dose equivalents as a covariate (all *P*>0.99 except L IFG self-inhibition, *P*>0.75), removal of the hallucinations score from the BPRS positive symptom total (all *P*>0.95 except L IFG and R A1 self-inhibition, *P*>0.75), and analysis without covariates (all *P*>0.99 except L IFG self-inhibition, *P*>0.75).

PEB analysis associated Digit Symbol score with decreased R IFG self-inhibition in Con (*P*>0.99; Figure 5E) but not in Scz, who instead showed decreased L A1-STG and increased L STG-A1 connectivity (both *P*>0.99, not shown).

We repeated the rsfMRI analyses without global signal regression (Figures S7A-E). Many results changed, with much greater between subject variability in parameters (the Scz > Con contrast, Figure S7A) or the loss of significant effects (Figures S7B, D and E).

### Self-inhibition findings across EEG and rsfMRI paradigms are similar but individual parameter fitting must be improved for use as a model-based biomarker

In summary, we found clear evidence for increased self-inhibition (evidence of reduced synaptic gain) in Scz (Figure 6A) in all data modalities and paradigms. However, *disinhibition* within auditory areas was associated with auditory perceptual abnormalities within Scz (Figure 6B). Furthermore, reduced self-inhibition was associated with Digit Symbol performance, especially in Con (Figure 6C). Note that associations with independent data speak to the predictive validity of the synaptic biomarkers furnished by DCM. A sensitivity analysis (see Online Methods) confirmed that increased superficial pyramidal self-inhibition best reproduced the key data features of the MMN (i.e. decreased MMN amplitude but unchanged latency; Figure S8A) and – along with loss of sp-ii connectivity – the decreased 40 Hz ASSR (Figure S8B).

**Figure 6.**
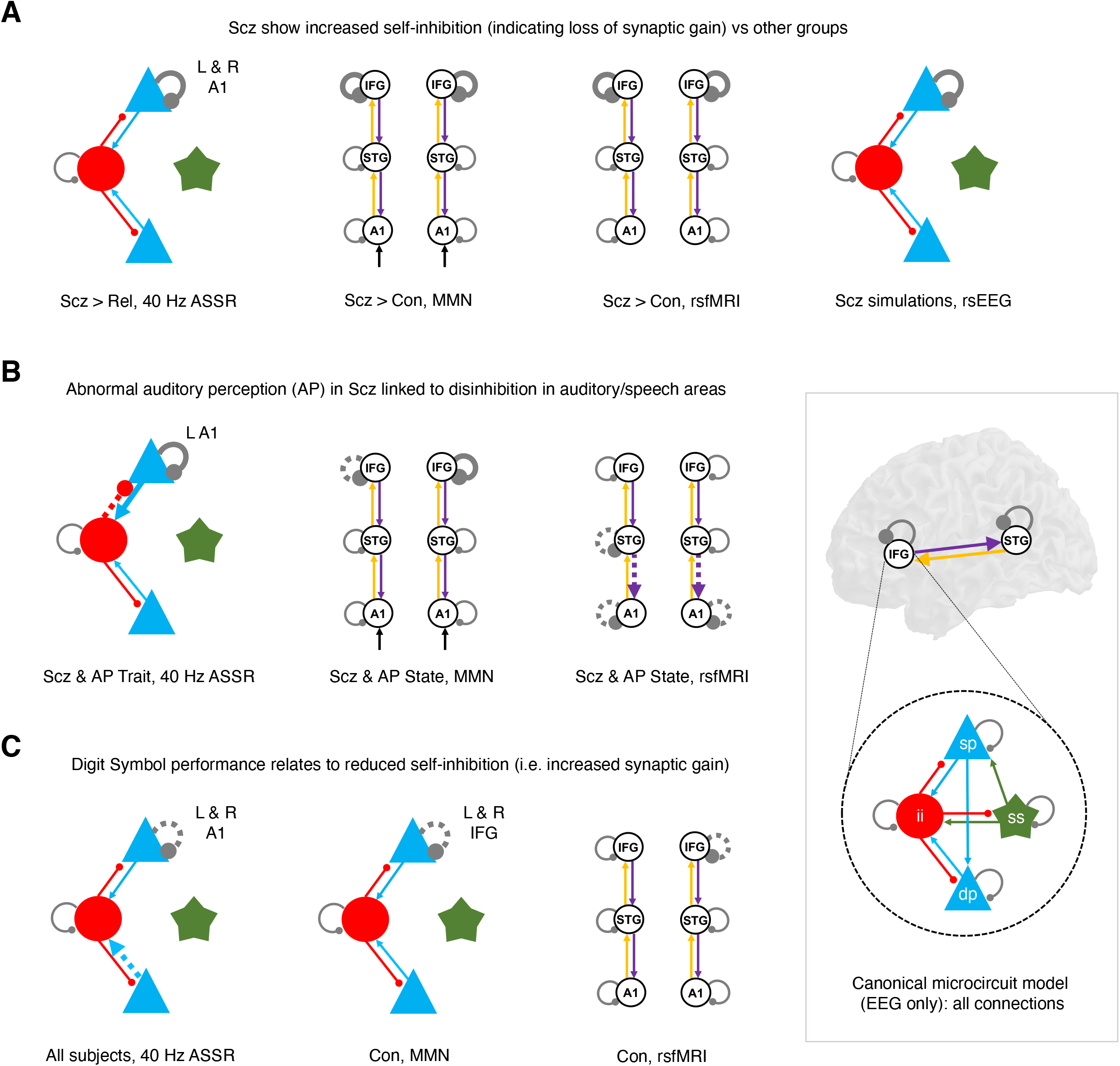
Summary of key findings across paradigms. This figure illustrates similar DCM findings across paradigms using the schematic illustrations from previous analyses. The inset at bottom right shows the canonical microcircuit model for EEG (below), which exists in each modelled cortical area (above). The microcircuit consists of superficial and deep pyramidal cells (sp and dp, blue), inhibitory interneurons (ii, red), and spiny stellate cells (ss, green), interconnected with excitatory (arrowheads) and inhibitory (beads) connections. A – Crucially, the Scz group consistently exhibited increased self-inhibition (as expected from a loss of synaptic gain) – in superficial pyramidal cells in particular (i.e. in the EEG paradigms). This was the case (from left to right) in A1 in the 40 Hz ASSR (when compared with Rel), in bilateral IFG in both the MMN (deviant–standard contrast) and the resting state fMRI, and in the rsEEG simulations. B – Within the Scz group, abnormal auditory percepts were linked with disinhibition in A1 in both the 40 Hz ASSR paradigm and the resting state fMRI, and with disinhibition in L IFG – i.e. Broca’s area – in the MMN (deviant–standard contrast). C – There is a consistent relationship of Digit Symbol task performance to reduced self-inhibition (i.e. increased synaptic gain) in Con in the MMN and resting state fMRI, and across all participants in the 40 Hz ASSR paradigm.

We also assessed whether the synaptic differences in Scz might be driven by early or late illness stages. We did not have access to illness duration data, but instead repeated the Scz > Con group comparisons for each paradigm, after dividing the groups equally into younger (≤36 years) and older (≥37 years) participants. The group differences in rsEEG features, MMN parameters and 40 Hz ASSR parameters were driven by the older group (Figure S9A-C); the rsfMRI parameter differences by the younger group (Figure S9D).

Finally, in *post hoc* analyses, we asked whether self-inhibition parameters – that were altered in Scz – had predictive validity across paradigms, thus licensing their use as ‘model-based biomarkers’. We used PEB to assess relationships in Scz between self-inhibition parameters in IFG in the MMN, or in A1 in the 40 Hz ASSR, and their corresponding self-inhibition parameters in the rsfMRI, in participants whose EEG and rsfMRI were <100 days apart (MMN n=44, 40Hz ASSR n=40), as some participants underwent EEG and rsfMRI many months or even years apart. Within Scz, there was evidence of associations between self-inhibition in R IFG in the MMN and rsfMRI (*P*>0.95; Figure S10A) and in R A1 in the 40 Hz ASSR and rsfMRI (*P*>0.95; Figure S10B). However, there were no such relationships in the left hemisphere or in Con.

To explore these relationships further we assessed their Pearson correlations (Figure S10C). None were significant (all *P*>0.05, uncorrected), and posterior parameter estimates clustered around the priors (of 0 in the 40 Hz ASSR, and 0.5 in the MMN), especially in the MMN, indicating the PEB results were driven by the subset of parameters estimated with sufficient efficiency to be drawn away from the prior.

## Discussion

Dynamic causal modelling of EEG and fMRI produced several key cross-paradigm findings. First, well-established effects in rsEEG^27^, MMN^11^ and 40 Hz ASSR^12^ paradigms in Scz were replicated and all could be explained by increased self-inhibition in (superficial) pyramidal cells. Likewise, Scz also showed an increase in prefrontal self-inhibition – as in the MMN – in rsfMRI (Figure 6A). Given the pathophysiology of Scz, this most likely reflects diminished synaptic gain on pyramidal cells^28^ (discussed below).

Second, abnormal auditory percepts in Scz was associated with *decreased* self-inhibition in auditory areas selectively, across three paradigms (Figure 6B). This is consistent with 40 Hz ASSR γ power^29^ (and phase locking of auditory γ^30^) correlating positively with auditory symptoms, despite being reduced in Scz overall (as in the visual domain^31^). BPRS positive symptoms were also associated with disinhibition in the rsfMRI analysis (Figure 5D). These apparently opposing effects of group and symptoms on self-inhibition can be explained by current models of Scz^1,4^ whereby decreased synaptic gain (NMDAR hypofunction in particular) is compensated by allostatic disinhibition of pyramidal cells (i.e. interneuron downregulation). These results suggest that psychotic symptoms are the price paid for this rebalancing of excitatory and inhibitory transmission.

Third, processing speed (i.e. Digit Symbol score) in Con was associated with disinhibition across three paradigms (Figure 6C). This plausibly reflects a relationship between cognitive performance and synaptic gain, and complements a recent rsfMRI DCM study of healthy adults (n=602) that found fluid intelligence was related to decreased self-inhibition in dorsal attention and salience networks^32^. In Scz, self-inhibition and cognition were only associated – in the 40 Hz ASSR – in R A1. Interestingly, a rsEEG DCM study in Down syndrome (n=36) found intelligence was associated with self-inhibition in V1^33^. Why is unclear: self-inhibition may be estimated more efficiently in sensory regions.

Regarding the rsEEG, increased θ power in Scz rsEEG is a well-established finding^27,34^, as is a ‘U-shaped’ change in spectral power (i.e. increased θ, decreased β, increased γ), although this pattern has been seen across θ, α and β frequencies^27^. Surprisingly, older Scz drove the increased γ effect: low γ (30-45 Hz) power is typically reduced in Scz with longstanding diagnoses^35^. Age is an imperfect proxy for illness duration, however.

The decreased mismatch effect in Scz (and especially in chronic Scz^36^) is well-documented^11^, and we found an effect of similar size in Rel – larger than is typical^36^. Underlying this effect, we found oddball stimuli decrease self-inhibition in IFG in Con, but not in Scz: recapitulating other DCM studies of Scz^13,26^. The mismatch effect rarely correlates with hallucinations in Scz (e.g. in only 3/22 studies^11^), but we found abnormal auditory percepts related to (condition-specific) disinhibition in L IFG – Broca’s area. Traditional MMN analysis (using electrode Fz) might miss this lateralised effect. Nevertheless, there are reports of left-lateralised associations of hallucinations (including IFG) with auditory oddball responses in Scz^37^.

In the 40 Hz ASSR paradigm, Scz showed decreased γ power and peak frequency, and Rel decreased power (as elsewhere^12,24,38^). DCM indicated that diminished pyramidal connectivity to interneurons was common to both Scz and Rel but loss of pyramidal gain was unique to Scz (Figure 4G). Others have modelled 40 Hz ASSR in Scz by increasing interneuron time constants^39^: this reproduced a concurrent increase in 20 Hz power in Scz^39^, which was not observed in our data. We assumed time constants did not differ in Scz in the ASSR or MMN, and estimated connectivity parameters – and delays, in the ASSR – instead (these can be regarded as synaptic rate constants).

A previous rsfMRI DCM analysis in Scz found disinhibition in anterior cingulate cortex^25^, rather than increased self-inhibition in bilateral IFG (Figure 5A). This combination speaks to a pattern of altered intra-prefrontal functional connectivity in early Scz^40^: namely, increased connectivity of medial areas and more modest decreases in connectivity in lateral areas. Prefrontal hyperconnectivity correlated positively with positive symptoms^40^. We similarly found positive symptoms were associated with disinhibition in bilateral IFG, and also A1 (Figure 5D). This relationship echoes findings that increased functional connectivity of primary sensory areas (to thalamus) correlates with PANSS scores^41^, and that increased A1 rsfMRI autocorrelation (a result of reduced self-inhibition) in Scz relates to auditory hallucinations^42^ (c.f. Figure 5C, right). R IFG self-inhibition’s relationship to Digit Symbol score (in Con; Figure 5E) mirrors the finding that the global functional connectivity of a nearby region of left lateral prefrontal cortex relates to fluid intelligence^43^. Taken together, these results may indicate that some functional connectivity relationships to either symptoms or cognition may not depend on extrinsic connections between nodes, but on synaptic gain within nodes.

Although global signal regression removes physiological ‘noise’, the global signal has greater power in Scz, which may have a neuronal component^44^. Here, GSR strengthened existing but weak effects in both Scz and Con (Figures 5C-E and S7C-E), and converted reduced forward connectivity into IFG (amidst a large amount of between-subject parameter variability, Figure S7A) to reduced synaptic gain within IFG. Given this, and given GSR in Scz affords a relatively uniform transform of the data^44^, the results with GSR are probably more reliable, especially given their consilience with the EEG results (Figure 6).

The Rel group showed mixed effects across paradigms, and more data are required to draw firm conclusions. In the MMN, no effects exceeded *P*>0.95 despite Rel’s similar data features to Scz, perhaps because the Rel group was smaller. In the 40 Hz ASSR, pyramidal self-inhibition was reduced in Rel (Figure S5B), not increased (as in Scz). In the rsfMRI however, Rel showed comparable IFG self-inhibition increases relative to Scz (Figure 5B). Interestingly, Rel’s pattern of apparently normal (or decreased) self-inhibition in the EEG paradigms – yet increased self-inhibition in rsfMRI – was recapitulated in the younger Scz (Figure S9).

A crucial question is what changes in ‘self-inhibition’ mean: changes in synaptic gain, or reciprocal coupling with interneurons? Our interpretation of self-inhibition changes is guided by known pathophysiology in Scz: i.e. given cortical synaptic gain is decreased (e.g. reduced function of NMDA^1,2,5^, dopamine 1^9^ and muscarinic^10^ receptors), and inhibitory interneurons downregulated^4,7^, then the simplest interpretation of increases and decreases in pyramidal self-inhibition are diminished pyramidal synaptic gain^28,45^ and decreased interneuron function, respectively.

Regarding potential causes of reduced synaptic gain, some Scz data features imply NMDAR hypofunction. In rsEEG, increased γ follows NMDAR antagonism^46^, e.g. using ketamine (which also suppresses β)^47^ or in NMDAR encephalitis (which also increases θ)^17,48^. In contrast, LSD and psilocybin do not increase θ^49^, and dopamine 2 antagonists potentiate α and β^50,51^. The 40 Hz ASSR is sensitive to NMDAR function^20^ (but also cholinergic^52^, dopaminergic^53^ and serotonergic^54^ manipulations): the latter do not affect the MMN, however, which is quite specific to NMDAR function^11^. Ketamine also reduces rsfMRI functional connectivity of IFG and auditory cortices^55^. Antipsychotic dose covariates weakened the Scz MMN condition-specific effects (Figure 3F) but strengthened the overall MMN effects (Figure S4C); they also weakened the Scz rsfMRI effects, but similar rsfMRI effects emerged in unmedicated Rel (Figure 5). Overall, these findings resemble NMDAR hypofunction, and seem unlikely to be medication-induced.

Several limitations are addressable: given pathophysiology is dynamic in Scz^1^, and that subgroups may exist^56^, larger datasets should be analysed, containing more early course (and preferably unmedicated) Scz. Furthermore, we must understand why younger Scz drive rsfMRI self-inhibition effects yet older Scz drive EEG self-inhibition effects. DCM models with explicitly parameterised NMDA (and other) receptor conductances^17^ can explore ‘self-inhibition’ in more detail. Our analyses were restricted to a few cortical regions, to maximise cross-paradigm effects, but future work should broaden this focus, especially to other frontotemporal regions.

Another key limitation involved the efficiency of EEG model inversion. We found preliminary evidence for a relationship between self-inhibition parameters across EEG and fMRI paradigms in Scz (Figure S10), but the clustering of EEG estimates around their prior values (Figure S10C) must be finessed (by using more informative data or paradigm design) to assess cross-paradigm self-inhibition reliability in all participants. If parameters show cross-paradigm validity, hierarchical PEB can be used to estimate them from multiple paradigms simultaneously^21^.

In conclusion, we found that dynamic causal modelling of multimodal neuroimaging data in Scz produced some remarkably consistent results across several paradigms. These include increased self-inhibition (i.e. diminished synaptic gain) in Scz, especially in frontal areas, but disinhibition – in auditory areas in particular – correlating with psychotic symptoms and auditory perceptual abnormalities specifically. Psychotic symptoms may therefore be caused by the interneuron downregulation that is necessary to restore cortical ‘excitation/inhibition balance’ in Scz. With appropriate model-fitting procedures, and larger datasets, model-based biomarkers for psychotic disorders may be at hand.

## Data Availability

Data are available upon reasonable request to Prof Elliot Hong.

## Acknowledgements

We are very grateful to numerous colleagues who provided guidance in DCM, including Vladimir Litvak, Peter Zeidman, Rosalyn Moran, Adeel Razi, Amirhossein Jafarian and Marta Garrido. RAA is an MRC Skills Development Fellow (MR/S007806/1) and has been supported by a UCL Bogue Research Fellowship, the Academy of Medical Sciences (AMS-SGCL13-Adams), the National Institute of Health Research (CL-2013-18-003), and the NIHR UCLH Biomedical Research Centre. JDM is funded by NIH grant MH112746. KJF is funded by the Wellcome Trust (Ref: 088130/Z/09/Z).

## Author contributions

RAA conceived the project, conducted or supervised all analyses, and wrote the paper. DP and KT developed analysis code and conducted some analyses. LU, AM, AMH, LC and JLJ assisted with analysis. GR developed analysis code and assisted with analysis. JDM contributed to the paper. KJF developed analysis code, assisted with analysis, and contributed to the paper. LEH collected the data and contributed to the paper. AA supervised the project and contributed to the paper.

## Online Methods

### Participant characteristics and measures

Data were collected from studies sponsored by the National Institute of Health (MH085646 and DA027680) and under human subject protocols approved by the University of Maryland, Baltimore, Institutional Review Board (HP-00045716 and HP-00043082) and University College London Research Ethics Committee (4356/003). All participants gave written informed consent prior to participation. Data were collected between December 2010 and April 2016. The study participants comprised those with a diagnosis of schizophrenia (Scz, n=107), controls who were approximately matched for age, gender and smoking status (n=108), and first degree relatives of Scz (n=57): details of the groups are listed in Table S1. Diagnoses were made using the Structured Clinical Interview for DSM Diagnoses (SCID)^1^. Scz were recruited from outpatient clinics, relatives from Scz participants themselves and also from media advertisements, and controls from media advertisements. Exclusion criteria were: neurological illness, head injury, substance abuse or dependence (except for nicotine), and daily use of benzodiazepine drugs. Controls and relatives were excluded if they met criteria for an Axis I diagnosis using the SCID, and controls were required to have no family history of psychosis within two generations.

All participants completed the Digit Symbol Substitution Task^2^ – a symbol-copying task that assesses learning, executive function and processing speed – and the Auditory Perceptual Trait and State scale (APTS)^3^. The APTS is a self-rated scale that assesses the frequency of abnormal auditory percepts, from altered characteristics of sounds to illusions, pseudohallucinations and verbal and non-verbal hallucinations. The scale includes ‘trait’ and ‘state’ measures, defined as symptoms experienced over one’s lifetime until two weeks ago, and over the past week, respectively. The full scale is available at http://www.mdbrain.org/APTS.pdf. Its test–retest reliability was assessed in 41 participants about 4 months apart, which showed ICC = 0.81 for both the trait and state measures, suggesting good reliability.

Use of psychotropic medication was recorded. 90 Scz were taking one or more antipsychotics (including Clozapine; see Table S1), for 15 Scz no medication was recorded and 3 were unmedicated. Some participants were also taking antidepressants, hypnotics (as prescribed) or mood stabilisers – the Scz group had higher proportions in each case. The 20 item Brief Psychiatric Rating Scale (BPRS) was used to rate overall symptoms in the Scz group only.

### Data acquisition and paradigms

For EEG recording, participants sat in a semi-reclining chair inside a sound-attenuated chamber, wearing an electrode cap. 64 scalp electrode sites were recorded, according to the 10/20 International System, using Neuroscan Stim acquisition software and a Synamps2 or Synamps2 RT amplifier. Recordings were grounded midway between FPZ and FZ using silver/silver-chloride electrodes and referenced to the nose. Eye movements were monitored by vertical and horizontal electrooculograms (EOGs). Data were continuously sampled at 1000 Hz with a DC/100 Hz band-pass filter (24 dB/octave roll-off). Impedances were kept below 5 kΩ.

For the resting state EEG (rsEEG) paradigm, subjects were asked to remain awake whilst EEG data were recorded. Two recordings were obtained on the same day, one with eyes open, and one with eyes closed. Each recording lasted 5 minutes.

In the mismatch negativity (MMN) paradigm, participants were presented with 1000 auditory stimuli through earphones, of which 800 (80%) were standard tones presented at 75 dB, for 60 msec, at 1000 Hz; 200 (20%) were duration-deviant tones at 75 dB, for 150 msec, at 1000 Hz. All tones had a 5 msec rise/fall time, with a stimulus onset asynchrony of 300 msec. 800 tones were presented in a single block. Participants were asked to ignore the tones while viewing a silent movie. Some of the MMN data have been analysed elsewhere^4^, using classical statistical tests and structural equation modeling to find correlations between MMN responses and neurotransmitter levels. Here we obtained effective connectivity estimates using DCM.

In the auditory steady state response (ASSR) paradigm, participants listened to click trains delivered by headphones, at 2.5, 5, 10, 20, 40, and 80 Hz. 75 stimulus trains (trials) – each consisting of 15 clicks, with each click at 72 dB and of 1 ms duration – were delivered at each stimulus frequency. The duration ranged from 6 s per train for 2.5 Hz, to 0.1875 s per train for 80 Hz. The inter-train interval was 0.7 s. Therefore, the durations for 2.5, 5, 10, 20, 40, and 80 Hz were 8.38, 4.69, 2.82, 1.89, 1.42, and 1.19 min, respectively, presented in 6 separate blocks separated by 2 min. The order of the blocks was randomized. In this study, only the data for the 40 Hz ASSR were analysed: data from this and the other frequency bands have been analysed elsewhere^3^.

For the resting state fMRI (rsfMRI) paradigm, participants were asked to keep their eyes closed and relax. fMRI data were collected at the University of Maryland Center for Brain Imaging Research using a Siemens 3T TRIO MRI (Erlangen, Germany) system equipped with a 32-channel phase array head coil. Resting state functional Blood Oxygen Level Dependent (BOLD) T2*-weighted images were obtained with axial slices parallel to the anterior-posterior commissure (AC-PC) using a T2*-weighted gradient-echo, echo-planar sequence (TR=2.2 s, TE=2.8 ms, flip angle=13°, FOV=256 mm, 128×128 matrix, 1.94×1.94 in-plane resolution, 4mm slice thickness, 37 axial slices, 444 volumes per run; 1 run). Acquisition lasted around 16 minutes. Structural images were acquired using a T1-weighted, 3D magnetization-prepared rapid gradient-echo (MPRAGE) sequence (TR/TE/TI=2200/4.13/766 ms, flip angle=13°, voxel size [isotropic]=0.8 mm, image size=240×320×208 voxels), with axial slices parallel to the AC-PC line.

### Data preprocessing

All EEG preprocessing and analysis was performed within Matlab (The MathWorks Inc.). MMN and 40 Hz ASSR data were imported into EEGLAB^5^ (https://sccn.ucsd.edu/eeglab/) and band-pass filtered between 1 and 70 Hz, with a notch filter at 59.5-60.5 Hz. rsEEG data were band-pass filtered between 1 and 50 Hz as higher γ frequencies are more vulnerable to artefacts in non-timelocked data. The data were then epoched: rsEEG into 6 s epochs, MMN into epochs from −50 ms to 300 ms around the stimulus onset, and 40 Hz ASSR data into epochs from −100 ms to 600 ms around the stimulus onset. The data then underwent an automated artefact rejection pipeline within EEGLAB, also employing the Multiple Artefact Rejection Algorithm (MARA)^6^. First, epochs with amplitudes of >±5 std were rejected, as were epochs containing linear trends (using pop_rejtrend, max slope=5, min R2=0.7) and low (0-2 Hz) or high (20-40 Hz) frequency bands (using pop_rejspec) within power thresholds of −50 to 50 dB and −100 to 25 dB respectively (40 Hz ASSR data used a frequency band of 20-35 Hz given the click train was presented at 40 Hz). Channels were then rejected (using pop_rejchan) on the basis of extreme values in their spectrum (−4 to 6 std), kurtosis (−7 to 15 std) or joint probability (−9 to 7 std). If >50% epochs were rejected, channel rejection was performed first, and epoch rejection second. If >50% epochs were still rejected, or if >20% channels were rejected, the data was discarded. Independent component analysis (ICA) was then performed, and components with p>0.5 of being artefacts – determined using stored artefact templates within MARA – were then rejected. MARA contains a supervised machine learning algorithm that learns from expert ratings of 1290 components by extracting features from the spectral, spatial and temporal domains, in order to identify artefacts of all kinds (e.g. eye, muscle, loose electrodes, etc). After MARA, channel and epoch rejection were run a second time using lower thresholds, as sometimes artefacts became apparent following component rejection. Channels were rejected on the basis of the following values in their spectrum (−6 to 5 std), kurtosis (−6 to 9 std) or joint probability (−7 to 7 std). Missing channels were then interpolated and the data were rereferenced to the common average. The data were then converted to SPM format and imported into SPM12 (v7219)^7^, available at https://www.fil.ion.ucl.ac.uk/spm/software/spm12/. The MMN and 40 Hz ASSR data were baseline corrected to the first 50 ms of the epoch, and the MMN data were downsampled to 250 Hz. The MMN (standard and deviant conditions) and 40 Hz ASSR were then averaged within each subject using robust averaging.

The structural MRI and rsfMRI data were preprocessed in accordance with the Human Connectome Project (HCP) Minimal Preprocessing Pipelines (MPP), adapted to be compatible with “legacy” data (single-band BOLD, no field map or T2-weighted image). These modifications to the MPP are described in detail in prior publications^8^ and are offered as a standard HCP MPP option at https://github.com/Washington-University/HCPpipelines/pull/156. Briefly, the T1-weighted images were first aligned in the volume space to the standard Montreal Neurological Institute-152 (MNI-152) brain template via a single-step combined transformation using the FMRIB Software Library (FSL) linear image registration tool (FLIRT) and non-linear image registration tool (FNIRT)^9^. FreeSurfer (http://surfer.nmr.mgh.harvard.edu/) was then used to segment gray and white matter in the whole brain to produce individual cortical-subcortical anatomical segmentations^10^ – specifically the pial and white matter cortical boundaries which were used to define a “cortical ribbon”, and a subcortical grey matter voxel mask. These were then used to define the hybrid surface/volume neural file for each subject in the Connectivity Informatics Technology Initiative (CIFTI) “grayordinate” space^11^.

BOLD images were first slice time corrected using FSL slicetimer. Next, motion correction was performed by aligning all BOLD data to the first frame of every run via McFLIRT. Motion corrected BOLD images were then registered to T1w image using FreeSurfer BBR. Finally, all the linear transformation matrices and nonlinear warp images were combined and the original slice time corrected BOLD images were registered to MNI-152 brain template in a single step. To exclude signal from non-brain tissue a brainmask was applied and cortical BOLD data were converted to the CIFTI gray matter matrix by sampling from the anatomically-defined cortical ribbon. These cortical data were then aligned to the HCP atlas using surface-based nonlinear deformation^11^. Subcortical voxels in the BOLD data were extracted using the Freesurfer-defined segmentation to isolate the subcortical volume portion of the CIFTI space.

In addition to the HCP MPP, all BOLD images had to pass stringent quality assurance criteria as previously reported^12^ to ensure that all functional data were of comparable and high quality: i) signal-to-noise ratios (SNR) >90, computed by obtaining the mean signal and standard deviation (sd) for a given slice across the BOLD run, while excluding all non-brain voxels across all frames^13^; ii) movement scrubbing as recommended by Power et al^14,15^. ‘Movement scrubbing’ refers to the practice of removing BOLD volumes that have been flagged for high motion, in order to minimize movement artefacts, and is a widely used fMRI preprocessing technique. Specifically, to further remove head motion artefacts, as accomplished previously^12^, all image frames with possible movement-induced artefactual fluctuations in intensity were identified via two criteria: First, frames in which sum of the displacement across all 6 rigid body movement correction parameters exceeded 0.5 mm (assuming 50 mm cortical sphere radius) were identified; second, root mean square (RMS) of differences in intensity between the current and preceding frame was computed across all voxels divided by mean intensity and normalized to time series median. Frames in which normalized RMS exceeded 1.6 times the median across scans were identified. The frames flagged by either criterion were marked for exclusion (logical or), as well as the one preceding and two frames following the flagged frame. Movement scrubbing was performed for all reported analyses across all subjects. Subjects with more than 50% of frames flagged were removed from subsequent analyses. Next, to remove spurious signal in resting state data we completed additional preprocessing steps, as is standard practice^16^: all BOLD time-series underwent high pass temporal filtering (>0.008 Hz), removal of nuisance signal extracted from anatomically-defined ventricles, white matter, and the remaining brain voxels (i.e. global signal) (all identified via participant-specific FreeSurfer segmentations^17^), as well as 6 rigid-body motion correction parameters, and their first derivatives using previously validated in-house Matlab tools^18^. Note that while the removal of global signal is a debated topic in fMRI^19,20^, it remains the gold standard for removing spatially pervasive artefact in the brain (although other techniques are emerging^21,22^). BOLD signal within the subject-specific cortical mask was spatially smoothed with a 6 mm full-width-at-half-maximum (FWHM) Gaussian kernel and dilated by two voxels (6 mm) to account for individual differences in anatomy, as done previously^13,23^. The cortical data were then parcellated using the 360-area multi-modal parcellation from Glasser et al. (2016)^24^.

### Data analysis and modelling: resting state EEG

The power spectrum for each electrode in each subject was computed using the default settings of Welch’s method in Matlab (6 s windows with overlaps of 3 s). Each power spectrum was normalised (sensor by sensor) by subtracting the 1/f slope in log space: this gradient was determined using the default settings (bisquare weight function) of robustfit in Matlab. The normalised spectra were averaged over all sensors and all subjects in each group. Group differences in normalised power in each frequency band were computed using the following definitions: θ 3-7 Hz, α 8-14 Hz, β 16-30 Hz, γ 31-50 Hz.

The biophysical model used to simulate power spectra is a convolution-based neural mass model^25^ in DCM. It is a neural mass model in the sense that it treats ensemble neural activity as a point process (without spatial extension) and only first order statistics (means) of neural population activities influence each other. Convolution-based models (as opposed to conductance-based models, which model ion channels in cell membranes) convolve presynaptic inputs (firing rates) with synaptic response kernels to produce postsynaptic membrane potentials in a given neural population^26,27^. The parameters summarising the kernel are the maximum postsynaptic depolarisation (determined by receptor density, or ‘connectivity’) and membrane time constants. These parameters are listed as connectivity parameters G and time constants T in Figure S1A. The model then transforms the neural population’s membrane potential back into a firing rate using a sigmoid operator whose gain or slope is controlled by parameter S. This firing rate becomes the input for another population, determined by the network’s intrinsic and extrinsic connectivity. Delays in transmission within the microcircuit (intrinsic) and from one brain area to another (extrinsic) are parameterised by D. The microcircuit itself (Figure S1A) contains four neural populations: spiny stellate cells (ss), superficial pyramidal cells (sp), deep pyramidal cells (dp) and inhibitory interneurons (ii). Extrinsic connections follow known anatomical patterns^28^: ‘forward’ connections project from sp cells to ss and dp cells in the area above, and ‘backward’ connections project from dp cells to sp and ii cells in the area below. Given all of the above, the dynamic activity of all populations in the network can be computed for any given input (for evoked responses like the MMN) or as a filtered spectral response to endogenous neuronal fluctuations (a mixture of pink and white neuronal noise) as in the rsEEG^25^.

Most of the analyses in this paper concern group differences in G parameters, i.e. connectivity between neural microcircuit populations (e.g. sp to ii) or self-inhibition (e.g. of sp to sp). Self-inhibitory connections may parameterise one or more of several physiological effects: i) they control the responsiveness of a population to its inputs, as any mechanism controlling synaptic gain does, e.g. NMDAR function, but also classical neuromodulatory receptors such as cholinergic and dopaminergic receptors; ii) they may reflect the action of an inhibitory interneuron population in a circuit, e.g. from sp cells projecting to parvalbumin+ (PV^+^) interneurons and back to sp cells^29^; iii) in the case of interneurons, they may reflect autapses (self-synapses: common on PV^+^ cells^30^). A model cannot distinguish between these mechanisms unless they are explicitly encoded in the model itself; however, some interpretations are more or less likely given what we know of pathology in schizophrenia. For example, a finding of increased sp self-inhibition in Scz is more likely to be due to loss of synaptic gain in those cells than a strengthening of the sp-PV^+^-sp circuit, which most evidence implies is weakened in Scz^31^. The opposite is true for a finding of decreased sp self-inhibition in Scz: its most likely explanation would be a loss of ii inhibition of sp cells.

Similarly, the microcircuit model does not explicitly distinguish between PV^+^ and somatostatin+ (SST^+^) interneurons. SST^+^ interneurons are of interest as their cortical markers are just as (if not more) reduced in Scz as PV^+32^. That said, the dynamics of the sp self-inhibition connection in the model are faster than those of the sp-ii-sp circuit (see the parameters in Figure S1A), hence the model can potentially distinguish between faster (likely PV^+^) and slower (e.g. SST^+^) interactions between pyramidal cells and interneurons^29^. (Note however that the empirical priors used for the 40 Hz ASSR analysis accelerated the dynamics of the sp-ii-sp circuit, in order to model the 40 Hz peak.)

The microcircuit model used was the same as that used by Shaw et al^33^: we termed this spm_fx_cmc_2017. It makes a small adjustment to spm_fx_cmc_2014, in that it replaces a G connection from sp to ss with a connection from sp to dp: see Figure 2C and Figure S1A. We used this model because it is closer to known anatomy^28^ than previous versions, but not too complicated to fit to EEG data (e.g. more sophisticated models^34^ contain separate interneuron pools for sp and dp cells). One key aspect of the model is that it models cortical dynamics only, and hence can model the γ and β peaks generated by superficial and deep cortical layers (respectively)^35–37^, but it cannot reproduce an α peak without adding an additional (thalamic) input^38,39^. Given the absence of group differences in α in the rsEEG, and for simplicity, we did not model this frequency band (as in previous studies^33,40^).

To simulate the power spectra in controls and Scz, we used the SPM12 (v7219) function spm_induced_optimise, which computes transfer functions (i.e. representations of cortical dynamics in the spectral domain) for parameters in the biophysical models in DCM. The simulated spectra were normalised in exactly the same way as the empirical spectra, subtracting the 1/f gradient using robustfit in Matlab. The standard priors in DCM were used (Figure S1A).

We did not try to fit the microcircuit model to the empirical rsEEG data (although this is possible^40^) because the choice of sources in rsEEG data can be problematic. We instead used the microcircuit model to simulate the effects of various potential circuit pathologies in Scz, and compared the results to the pattern in the rsEEG. The circuit pathologies were based on reasonable hypotheses about microcircuit connectivity abnormalities in Scz (Figure 2D):

**Model 1** – a <30% loss in connectivity in all microcircuit connections, i.e. a global loss of synaptic efficacy.

**Model 2** – a <30% loss of connectivity to/from interneurons (‘to’ and ‘from’ are identical from the modelling point of view – their effects are the same).

**Model 3** – a <30% loss of interneuron self-inhibition, to model potential disinhibition of PV+ interneurons by other PV+ interneurons^30^.

**Model 4** – a <30% gain in interneuron self-inhibition, to model loss of synaptic gain (e.g. hypofunction of NMDARs) on interneurons.

**Model 5** – a <30% gain in superficial pyramidal self-inhibition, to model loss of synaptic gain (e.g. hypofunction of NMDARs) on sp cells.

### Data analysis and modelling: mismatch negativity

The MMN data were plotted (using electrode Fz, as is standard practice^41^) as group-averaged waveforms for the standard and deviant tones separately (Figure S2A) and as the traditional ‘difference’ waves, i.e. deviant-standard waves, illustrating the mismatch effect (Figure 3A). Group differences in these waveforms were assessed using *t*-tests at each timepoint using an α of *p*<0.05 (uncorrected for multiple comparisons), and are displayed on the figures.

To more robustly illustrate the mismatch effects and group differences, incorporating other electrodes and cluster-based correction for multiple comparisons, the MMN sensor-level data were analysed in SPM12, after smoothing using an 8×8×8 mm FWHM Gaussian kernel (Figures 3B, 3C and S2B).

The sources of the MMN response are well characterised, e.g. using conjoint EEG-fMRI studies^42^. These are bilateral sources in primary auditory cortex (A1), secondary auditory cortex (superior temporal gyrus, STG) and inferior frontal gyrus (IFG). To ensure these source locations were adequate for our modelling, we source localised the MMN data using the ‘minimum norm’ model in SPM12. Priors on source locations (in MNI coordinates) were [-42, −22, 7] – left A1, [46, −14, 8] – right A1, [-61, −32, 8] – left STG, [59, −25, 8] – right STG, [-46, 20, 8] – left IFG, [46, 20, 8] – right IFG, and sources were estimated to be within a 16 mm radius of these prior locations. The activity peaks across all subjects and conditions were close to the standard sources (Figure S2C). Note that this source reconstruction was not used for the DCM analysis, however: source reconstruction (using the same priors) was performed as part of DCM model inversion. Joint optimisation of source locations and model parameters performs better than separate optimisations because it suppresses posterior correlations between source location parameters and biophysical parameters such that they explain away different parts of the data.

Standard DCM analyses estimate subject-specific parameters in four microcircuit connections (G): sp-sp, ii-ii, ii-sp and ii-dp. We estimated two extra G parameters given the strong possibility of differences in connectivity from pyramidal cells to interneurons in Scz^31^, i.e. sp-ii and dp-ii. To avoid overparameterising the model, however, we fixed parameters elsewhere. First, we estimated whole group posterior means for synaptic delays (D) and time constants (T) and then used these as priors, fixing the D and T parameters to these values (Figure S1A) in most models. Insodoing, we are not claiming that D and T are unlikely to be altered in Scz. We are merely assuming that there are very likely to be differences in G parameters in Scz, and that we wish to give the model the best chance of detecting them by reducing its degrees of freedom elsewhere. Second, we constrained all estimated G parameters to be the same (within but not between subjects) in every cortical area except for sp-sp self-inhibition. Underlying this is the reasonable assumption that microcircuit function (and its pathology in Scz) may be similar across the brain. Nevertheless, NMDAR (and other receptor) subtypes are differentially distributed along a cortical hierarchical gradient^43^, so if for example certain subtypes – preferentially located in higher or lower hierarchical areas – are more affected in Scz^44^, the model could detect these spatial differences in sp-sp connectivity. Given we were applying a brain-wide constraint to most G parameters, we used the whole group posterior means (Figure S1A) as priors, to reduce model fitting problems.

In total, six microcircuit models were evaluated (shown in Figure 3D):

**Models 4Ga, 4Gb, 4Gc, 4Gd** – these four models estimated four microcircuit connectivity (G) parameters each, consisting of sp-sp and ii-ii self-inhibitory connections and then different combinations of two of the four connections between sp, ii and dp cells. Delays (D) and time constants (T) were fixed to their group posterior estimates (see Figure S1A).

**Model 6G** – this model estimated all six G parameters of interest, but fixed D and T to their group posterior estimates.

**Model 6G,D,T** – this model estimated all six G parameters of interest and also D and T (using their group posterior estimates as priors).

Each of these models used the same macroscopic structure (Figure 2C, right), i.e. forward and backward connections linking areas in adjacent hierarchical levels, and lateral connections linking bilateral areas at the same level (not shown). Note that only forward, backward and self-inhibitory connections could show *condition-specific* differences between groups, i.e. differences between standard and oddball event related potentials.

MMN model fitting was performed using the ‘spatial’ or ‘IMG’ forward model in DCM for evoked response potentials (ERPs). DCM fits the first up to eight modes of the prior predicted covariance in sensor space^45^ (Figure S3A). In practice, the first 3-4 modes usually capture most of the variance (as in Figure S3A, right), so R^2^ values generated for the MMN data are based only on the first four modes. A boundary elements head model^46^ was used to approximate the brain, cerebrospinal fluid, skull and scalp surfaces.

Fitting non-linear neural mass models to EEG data is an ill-posed problem, and can lead to difficulties in optimisation, such as models getting stuck in local optima. Empirical Bayes for DCM^47^ can circumvent this issue by performing model fitting iteratively, each time using the group level posteriors over parameters as priors for each subject’s parameters in the next model inversion, yielding more robust and efficient parameter estimates^48^. In practice, it substantially improved model fit: model inversions performed with and without it had R^2^ values of ∼0.7 and ∼0.6 respectively. Local optima can also be avoided by fitting one fully parameterised model and then pruning unnecessary parameters using Bayesian model reduction (see below), rather than fitting many models with reduced numbers of parameters which are more prone to local optima problems^49^.

Following model inversion, models were formally compared using Bayesian model selection^50^ (Figure 3E). We used the protected exceedance probability^51^ as the metric of success: i.e. the probability that any one model is more frequent than the others, above and beyond chance. The R^2^ for the first four modes of the winning model was also computed (Figure 3E). Group differences between Con and Scz or Rel in their R^2^ values were also assessed using ranksum tests (as the distributions were skewed; Figure S3C).

Group differences in parameters and relationships between parameters and other measures were analysed using Parametric Empirical Bayes (PEB; spm_dcm_peb)^49,52^. PEB is a hierarchical Bayesian version of a general linear model that can assess which DCM parameters differ between groups or covary with a continuous measure. One critical advantage of PEB (over performing classical statistical tests on parameters) is that it takes account of not just each parameter’s expectation but also its covariance: therefore parameters that cannot be estimated with high confidence (e.g. in models that fit less well) contribute less to the inference.

A second key aspect of the PEB procedure is the use of Bayesian model reduction^53^ and Bayesian model averaging^49^ (spm_dcm_peb_bmc). In essence, these steps prune away unnecessary parameters and then obtain posterior estimates for the remaining parameters by averaging over the remaining models. The Bayesian model reduction procedure allows hypotheses about model parameters to be formally tested. This is done by first defining a model space of different parameter groupings that may best describe the modelled effect. In the MMN analysis, two model spaces were used: the first asked which combinations of *extrinsic or self-inhibitory* connections could best account for the *mismatch* effect (Figure S4A, left), and the second asked which combinations of *intrinsic* (microcircuit) connections could best account for the *overall group differences* across conditions (Figure S4B, left). These model spaces were based on the assumption that microcircuit parameters would not differ between conditions, but messages passed between areas (or pyramidal gain in specific areas) may well do so. In Figure S4A the first model (column) is a ‘null’ model, the second contains only ‘forward’ connections, the third contains only ‘backward’ connections, etc. The whole model space consists of different combinations of forward, backward, and self-inhibitory connections in either temporal or frontal areas.

Following Bayesian model reduction, the posterior probabilities of these models accounting for the group difference effect (Scz > Con) are plotted in the middle. This shows that the Scz > Con effect is best described by Model 7, containing only the left and right IFG self-inhibitory connections. Instead of simply using the parameters estimated from the winning model (as there may not always be a clear winner), posterior parameter estimates are obtained by averaging over all models, weighted by their model evidence (Bayesian model averaging). Note that if there is no evidence for some models, parameters unique to those models will be redundant: these parameters are eliminated following Bayesian model reduction. Conversely, parameters that are present in all probable models are highly likely to explain the group difference effect. The posterior probabilities of parameters differing between groups are shown on the right (Figure S4A): only six parameters have non-zero probabilities of explaining the group difference effect, but only two of these are >0.95. These probabilities generate the Bayesian confidence intervals for the group difference plots (Figure 3F, in this case).

Figure S4B (left) shows the model space for the group differences in microcircuit parameters. The first six rows correspond to sp-sp self-inhibitory connections in areas from L A1 to R IFG; the next five rows containing white boxes correspond to connections constrained to be identical throughout the model. The (column-wise) model space consists of different combinations of connections to and from inhibitory interneurons, and of sp-sp self-inhibition at different levels of the model. The subsequent plots show the model and parameter posterior probabilities; the results of the analysis are below in Figure S4C (middle and right).

### Data analysis and modelling: 40 Hz auditory steady state response

The timecourses of the 40 Hz ASSR waveforms in electrode Fz are plotted in Figure 4A: ∼40 Hz oscillations are superimposed on more sustained baseline changes (known to occur during auditory click trains^54^). Scz and Rel response baselines diverge (*t*-tests at each timepoint) from Con around 250 ms, but we restricted our modelling to well-replicated group differences in ∼40 Hz power. Each subjects’ EEG data were transformed into measurements of phase and power in the frequency domain using a Morlet wavelet in spm_eeg_tf. For the Fz sensor-level analyses, the spectra were normalised in order to assess the power at 40 Hz relative to the background 1/f slope. For normalisation, we again subtracted the 1/f gradient in log space (computed using robustfit in Matlab applied to the 10-30 Hz and 50-70 Hz ranges, on a subject-specific basis): example gradients of group averaged data from electrode Fz are shown in Figure S2D. γ peak frequency was the frequency in which the maximum (normalised) power within the 35-45 Hz range occurred (plotted in Figure 4B). The unnormalised group averaged time-frequency plots are shown in Figure S2E, and the normalised plots in Figure 4C. Permutation testing assessed whether the number of observed significant *t*-tests (*P*<0.05) for the Con v Scz, Con v Rel and Scz v Rel comparisons across all timepoints in the 30-55 Hz range shown was likely due to chance. There were no group differences in normalised power at other frequencies (*t*-tests at each timepoint and frequency, not shown).

The 40 Hz ASSR paradigm is known to activate primary auditory cortex (Heschl’s gyrus), anterior to auditory evoked response sources^55,56^. To obtain good priors for source localisation, we performed source localisation on all subjects in SPM12 using a minimum norm approach and focussing purely on the 37-43 Hz frequency band. Across all subjects, this generated three peaks of activation, either in or near right Heschl’s gyrus and anterior to the MMN sources. The closest to Heschl’s gyrus (A1) was [50 −12 4] (Figure 4D), which was used (along with [-50 −12 4]) as the seed for reconstruction of virtual electrode data from a broader band (35-50 Hz) for modelling in DCM.

The neuronal model for DCM had to be adjusted in order to model the high amplitude ∼40 Hz peaks in the virtual electrode data from left and right A1 (dotted lines in Figure S3B). To fit these peaks we introduced a Gaussian ‘bump’ at 40 Hz (of width w≤4 Hz) into the background neuronal noise, to reflect the specific 40 Hz input (similar to a previous model^39^ using a 10 Hz bump added to neuronal noise to simulate a thalamic input). As in the MMN analysis, we estimated group mean values for T (time constants) and also S (the slope of the sigmoid activation function) and J (the contribution of neuronal populations to the EEG signal) in an initial model inversion, and then used these as empirical priors (Figure S1A). These changes meant that sp and ii time constants were slightly shorter, there was increased gain in the transformation from membrane potential to firing rates, and that ss cells (which receive the 40 Hz thalamic drive) made more contribution to the measured EEG signal (respectively). We fixed T to its priors because without being fixed, they often became biologically implausible. We did not fix D (delays) because these might have important effects on peak frequencies in this paradigm. As in the MMN analysis, we estimated six microcircuit (G) parameters but constrained all except sp-sp self-inhibition to be identical (within subjects) in A1 bilaterally. Last, we increased the precision of the data (hE), which was necessary to force the model to fit the unnatural peaks in the 35-50 Hz range. We attempted to fit a wider range of frequencies (15-50 Hz), but the model fitted the lower frequencies at the expense of the 40 Hz peak, ignoring the key data feature, so the narrower (γ-only) band of 35-50 Hz was chosen.

40 Hz ASSR model fitting was performed using the ‘LFP’ forward model in DCM for cross-spectral densities (CSD), without iterative fitting (because the increased data precision caused unrealistic parameter changes). Model comparison between the model with new priors for T, S and J and models with previous versions of each of these priors was performed, to ensure the new priors improved the model (Figure 4E, left). The comparison also included a model with a narrower 40 Hz bump (w=1 Hz). The first ten subjects’ data (dotted lines) and model fits (thick lines) for the best model are shown in Figure S3B. The model fits reasonably well – it often cannot capture the full amplitude of the ∼40 Hz peak, but usually has its peak at the right frequency: one exception is subject 0254 (bottom right), whose unusual peaks at 45 Hz are too far from 40 Hz for the model to accommodate. R^2^ values for the winning model are shown in Figure 4E (right), and group differences between Con and Scz or Rel in their R^2^ values were also assessed (Figure S3C). R^2^ values were more variable in the 40 Hz ASSR than the other paradigms, because in the 40 Hz ASSR the model was trying to fit a small data feature very precisely.

The model space used for the analysis of A1 microcircuit (G) parameters within PEB is shown in Figure S5A (first). The first two rows denote sp-sp self-inhibition on the left and right, and the next five rows containing white boxes denote the remaining microcircuit parameters which were identical bilaterally. The (column-wise) model space is constituted by different combinations of sp and ii self-inhibitory parameters and ii input and output parameters. Following Bayesian model reduction, no model of the group differences dominates (middle), however all models with any evidence for them contain sp-sp self-inhibition parameters, so the posterior probability (after Bayesian model averaging) that these parameters contribute to the group difference is very high (right).

### Data analysis and modelling: resting state fMRI

We wished to restrict the rsfMRI analysis to being directly comparable to the MMN and 40 Hz ASSR analyses. We therefore chose to analyse effective connectivity in exactly the same network involved in the MMN (and 40 Hz ASSR, in part), i.e. bilateral A1, STG and IFG. The sources used for the MMN lie within areas A1, A4 and 44 of the Glasser parcellation^24^ (Figure 1), so these were chosen to be the nodes of the network. We did not use subject-specific sources from the MMN source reconstruction, because the MMN sources are well-established, and if a subject’s EEG source reconstruction lay outside these areas, it is arguably more likely that their EEG source reconstruction was inaccurate than that a different cortical network activated in their MMN. Thus, using the same (individually parcellated) rsfMRI sources for each subject took account of individual variation in anatomy whilst ensuring the rsfMRI analysis was robust and reproducible.

For the DCM analysis we used the movement-scrubbed rsfMRI data with global signal regression. Although global signal likely contains neuronal signal that is relevant to schizophrenia^57^, we removed it as we wished to reduce global physiological confounds as much as possible. No subjects were excluded due to low SNR, although doing so (for the ten subjects with SNR<25) did not change any of the results. Model structure was assumed to be as it was in the MMN analysis (Figure 1), with forward and backward connections linking adjacent hierarchical levels only, and lateral connections (not shown) linking bilateral areas at the same level; forward, backward and self-inhibitory connections could differ between groups.

Spectral DCM for rsfMRI was used to estimate effective connectivity. Spectral DCM estimates the amplitude and exponent parameters of the low frequency (<0.1 Hz) cross spectra of endogenous neuronal fluctuations, and thus the covariance of the underlying hidden states (i.e. neuronal activity in each node)^58^. The advantage of this power-law model is that it is deterministic – i.e. less computationally demanding – whilst accommodating stochastic fluctuations in neural states. In this model, the units of connectivity are Hz, i.e. stronger connectivity produces a faster rate of responding in a target region. The dynamics of the neuronal model are mapped to the fMRI signal via a haemodynamic forward model^59^. No model comparison was performed prior to PEB analysis because we wished to match the same model structure used in the MMN analysis, for comparative purposes, and priors did not need to be adjusted to improve model fit. The R^2^ values (and their differences between groups) are shown in Figure S3C.

Again, PEB was used to analyse group differences in parameters. The model space for the Scz > Con analysis is shown in Figure S6A (left). It comprises different combinations of forward, backward and self-inhibitory connections. There is no clear winning model (middle) but all likely models contained bilateral IFG self-inhibitory connections, hence these connections are inferred to be highly likely to explain group differences (right).

For comparative purposes, in addition to the spectral DCM analysis, we performed a standard functional connectivity analysis (Figure S6B), i.e. computing the Pearson (zero lag) correlation between two parcellations’ timeseries. We also computed their (zero lag) covariance (Figure S6C), because comparing correlations in Con and Scz can be problematic as their variances may differ^57^, and the corresponding zero lag covariance in the neuronal states estimated by DCM (Figure S6D). To see whether spectral DCM self-inhibition parameters might be recapitulated in basic data features, we also assessed group differences in the standard deviation of BOLD signal in each node (Figure S6E), given that greater self-inhibition would be expected to relate to lower variance in a node^60^.

It is clear from Figures 5A and S6B-E that one cannot reliably associate effective connectivity with functional connectivity or BOLD fluctuations. Reduced functional connectivity between A1 and STG in Scz (Figure S6B and elsewhere^61^) is less robust if covariance is used instead (an issue highlighted previously^57^). Conversely, increased IFG self-inhibition in Scz (Figure 5A) is not apparent in the BOLD standard deviations in IFG (Figure S6E), although reduced BOLD fluctuations in Scz have been reported in other areas^60^. Measures of functional connectivity differ from effective connectivity in several important ways: i) spectral DCM also estimates haemodynamic and neuronal/measurement noise parameters – which may contribute to group differences in functional connectivity e.g. if one group is older^62^ or taking (antipsychotic) medication, or if one group moves more (respectively); ii) spectral DCM connectivity includes non-zero lags, which indicate direction of connectivity^58,63^; iii) and – most importantly – spectral DCM also estimates self-inhibition within cortical regions.

We also performed the same DCM and PEB analyses without global signal regression, to ascertain the effects of this preprocessing step on the results (Figure S7).

### Data analysis and modelling: parameter sensitivity and cross-paradigm analysis

We performed parameter sensitivity analyses on the six estimated microcircuit (G) parameters to address two potential weaknesses in the MMN and 40 Hz ASSR analyses. In the MMN analysis, constraining condition-specific (mismatch) effects to only sp-sp self-inhibition amongst the G parameters – along with extrinsic connections – might miss such effects in other G parameters. In the 40 Hz ASSR analysis, the data were afforded high precision during model fitting in order to force the model to fit the unnatural 40 Hz peak: this may lead to overfitting of other data features, however, and spurious results. We therefore selected two subjects from each paradigm with EEG responses typical of the group average. We simulated virtual electrode (‘LFP’) data from a single cortical area using spm_induced_optimise and either spm_gen_erp (for the MMN) or spm_csd_mtf (for the 40 Hz ASSR). In each case we used these subjects’ posterior G parameter estimates, varying each G parameter by ±30% in turn; in the MMN, we also fixed delays (D) and time constants (T) to the values used in Figure S1A. The simulated data are shown in Figure S8, with the key data features that the parameters showing group differences ought to explain circled in red.

We sought to investigate whether Scz with shorter or longer illness durations were driving the key Scz > Con group difference effects in each paradigm. We thus repeated the Scz > Con analyses from each paradigm, splitting each group by median age, as we did not have illness duration data. Group differences in rsEEG power spectra in subjects ≤36 years old (Con n=50, Scz n=49) and in subjects ≥37 years old (Con n=48, Scz n=46) are shown in Figure S9A; group differences in MMN mismatch parameter effects in subjects ≤36 years old (Con n=45, Scz n=47) and subjects ≥37 years old (Con n=48, Scz n=48) in Figure S9B; group differences in 40 Hz ASSR parameters in subjects ≤36 years old (Con n=47, Scz n=48) and in subjects ≥37 years old (Con n=46, Scz n=46) in Figure S9C; and group differences in rsfMRI parameters in subjects ≤36 years old (Con n=44, Scz n=34) and in subjects ≥37 years old (Con n=42, Scz n=37) in Figure S9D.

We used the PEB framework to assess whether the abnormal parameters in the Scz group related to each other across modalities, i.e. across EEG and fMRI. If so, this would support the use of multiple paradigms in combination to estimate these ‘biomarker’ parameters. We selected the left and right IFG self-inhibition parameters from the MMN and left and right A1 self-inhibition parameters from the 40 Hz ASSR, and their counterparts from the rsfMRI. For each of the four rsfMRI parameters separately, we used the PEB framework to reveal which of all MMN or 40 Hz ASSR parameters related to it. Thus the posterior expectation and covariance in the EEG parameters, but only the expected value of the rsfMRI parameter, was used by the PEB model. (We reasoned that the rsfMRI parameters were likely estimated with greater confidence, given their simpler models and more consistently high R^2^ values compared with the EEG models (Figure S3C), hence it would be better to include the EEG parameters’ covariance in the PEB model.) Two of the four analyses yielded significant results: R IFG self-inhibition across rsfMRI and MMN (Figure S10A), and R A1 self-inhibition across rsfMRI and 40 Hz ASSR (Figure S10B).

For comparative purposes, we also performed classical analyses of the relationships between the same rsfMRI and EEG parameters. The Pearson correlations between the rsfMRI and 40 Hz ASSR or MMN self-inhibition parameters are shown in Figure S10C. Both EEG paradigms – especially the MMN – show some clustering of the parameters around their prior values (0 in the 40 Hz ASSR, and 0.5 in the MMN). Posterior parameter values that are further from the priors are typically estimated with greater confidence. The advantage of the PEB framework is that it will (optimally) upweight the contribution of these more confident parameters to group-level inference.

