## Supplemental Figures and Table for "Computational modelling of EEG and fMRI paradigms reveals a consistent loss of pyramidal cell synaptic gain in schizophrenia"

#### Supplement

Supplementary Figure legends: S1-S10

Table S1

### Fig S1: DCM parameters for EEG paradigms, additional rsEEG results

**A**

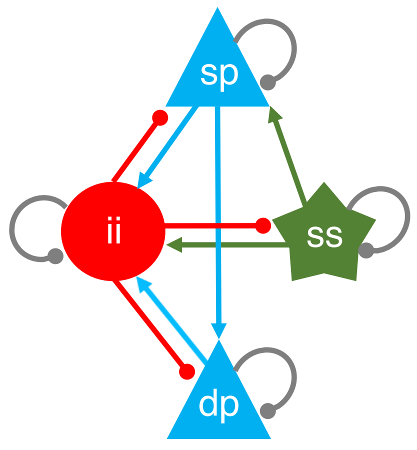

Full model
(all connections)

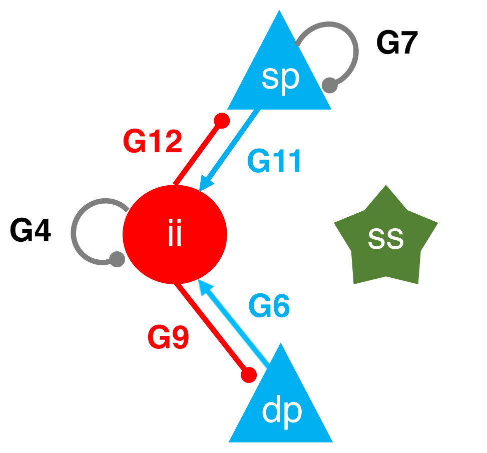

Connections permitted to differ between groups in 6G model

| Para meter | Description | rsEEG prior mean | MMN prior mean | MMN prior precision | 40 Hz ASSR prior mean | 40 Hz ASSR prior precision |
|---|---|---|---|---|---|---|
| G1 | ss self-inhibition (gain) | 800 | 800 | 0 | 800 | 0 |
| G3 | ii connectivity to ss | 800 | 800 | 0 | 800 | 0 |
| **G4** | ii self-inhibition (gain) | 800 | 240 | 1/32 | 800 | 1/32 |
| G5 | ss connectivity to ii | 800 | 800 | 0 | 800 | 0 |
| **G6** | dp connectivity to ii | 400 | 270 | 1/32 | 400 | 1/32 |
| **G7** | sp self-inhibition (gain) | 800 | 1300 | 1/32 | 800 | 1/32 |
| G8 | ss connectivity to sp | 800 | 800 | 0 | 800 | 0 |
| **G9** | ii connectivity to dp | 400 | 730 | 1/32 | 400 | 1/32 |
| G10 | dp self-inhibition (gain) | 200 | 200 | 0 | 200 | 0 |
| **G11** | sp connectivity to ii | 800 | 1800 | 1/32 | 800 | 1/32 |
| **G12** | ii connectivity to sp | 800 | 980 | 1/32 | 800 | 1/32 |
| G13 | sp connectivity to dp | 800 | 800 | 0 | 800 | 0 |
| T1 | Time constant: ss | 2 ms | 2 ms | 0 | 5 ms | 0 |
| T2 | Time constant: sp | 2 ms | 2.5 ms | 0 | 1.5 ms | 0 |
| T3 | Time constant: ii | 16 ms | 26 ms | 0 | 8 ms | 0 |
| T4 | Time constant: dp | 28 ms | 60 ms | 0 | 38 ms | 0 |
| D1 | Delay constant: intrinsic | 1 ms | 0.8 ms | 0 | 1 ms | 1/64 |
| D2 | Delay constant: extrinsic | 8 ms | 8 ms | 0 | 8 ms | 1/64 |
| S | Slope of sigmoid activation function | 1 | 1 | 1/64 | 1.6 | 1/64 |
| w | Maximum width of ~40 Hz Gaussian in spectrum | n/a | n/a | n/a | 4 Hz | 0 |
| J | Contributing states to MEEG [ss sp ii dp] | n/a | [0 1 0 0] | [1/32 0 0 1/32] | [0.3 1 0 0] | [1/32 0 0 1/32] |
| hE | Data precision | n/a | 6 | $0.5^5$ | 16 | $0.5^{14}$ |

**B**

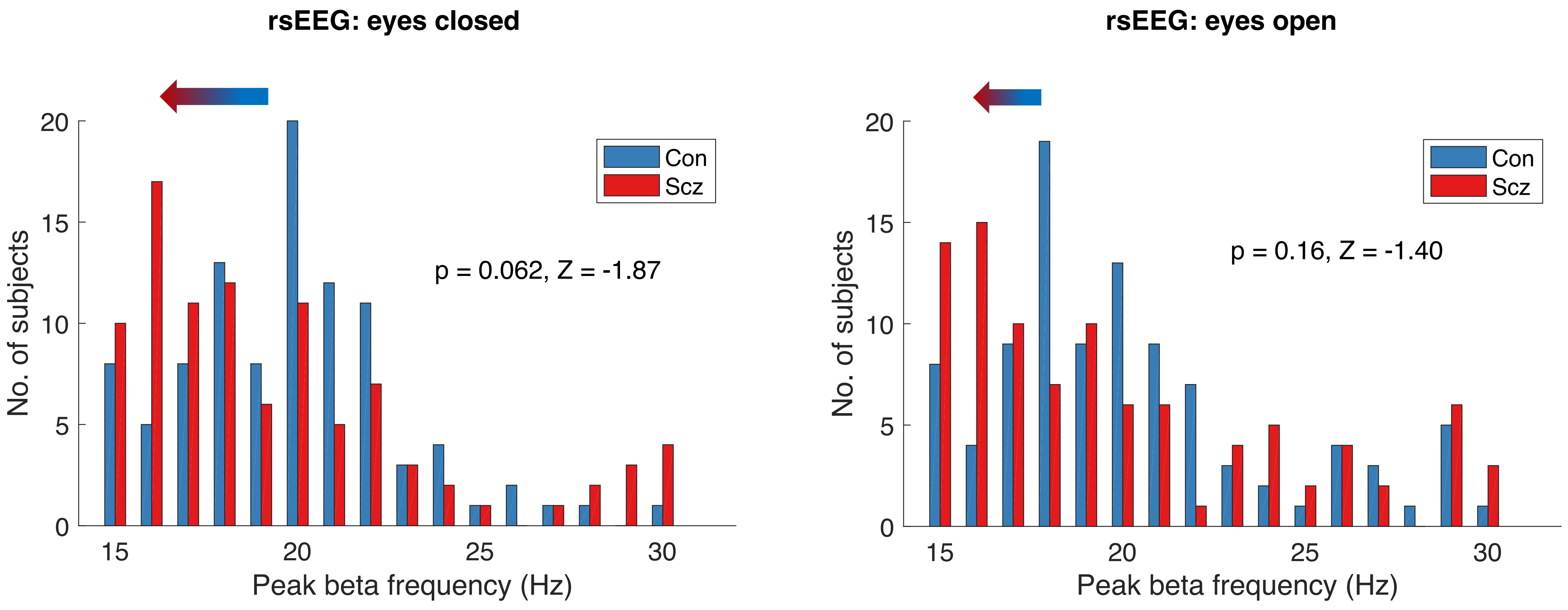

**Figure S1 – DCM parameters for EEG paradigms and additional resting state EEG results**

A – Left: The top panel shows the cortical microcircuit model used in the EEG paradigms. The four neuronal populations are spiny stellate cells (ss), superficial and deep pyramidal cells (sp and dp) and inhibitory interneurons (ii). Each send either excitatory or inhibitory connections to other neuronal populations (coloured connections: arrows are excitatory, balls are inhibitory), and also have self-inhibitory connections (grey) that parameterise their responsiveness to their input, i.e. synaptic gain. The bottom panel shows the subset of connections in this microcircuit that were estimated in the MMN and 40 Hz ASSR paradigms, and could thus differ between groups.

Right: The table contains the parameters (prior expectation and precision or inverse variance) of the EEG DCMs. All parameters except the final two rows pertain to the neuronal model; the final two pertain to the forward model (i.e. how the neuronal model generates the EEG signal). G denote 'intrinsic' connectivities, i.e. connection strengths within the microcircuit, in arbitrary units. T denote time constants in each neuronal population, and D denote transmission delays between populations (intrinsic) or between areas (extrinsic). w was a new parameter used to model the ~40 Hz peak in the power spectrum introduced by the click train in the 40 Hz ASSR paradigm. J, the states contributing to the EEG signal, assumes that sp cells make the biggest contribution, but also permits ss and dp cells to do so. DCM was only used to simulate rsEEG power spectra, hence no forward model parameters (or parameter precisions) were used. The neuronal model parameter settings were unchanged from their standard values. In the MMN analysis, group mean T and D parameters were estimated and then fixed to these values; six G parameters were also estimated and used as priors. Standard forward model settings were used. In the 40 Hz ASSR, T were again fixed to the group mean values, and group mean values were used as priors for S, w and J. The data precision was increased to ensure the model fitted the unnatural 40 Hz peak.

B – These histograms depict the numerical (but not significant) reduction in rsEEG peak β frequency in Scz (n=95) versus Con (n=98): in eyes closed (left) and open (right) conditions. The *P* values denote ranksum tests (not corrected for both comparisons).

### Fig S2: Additional MMN & 40 Hz ASSR (not normalized) results

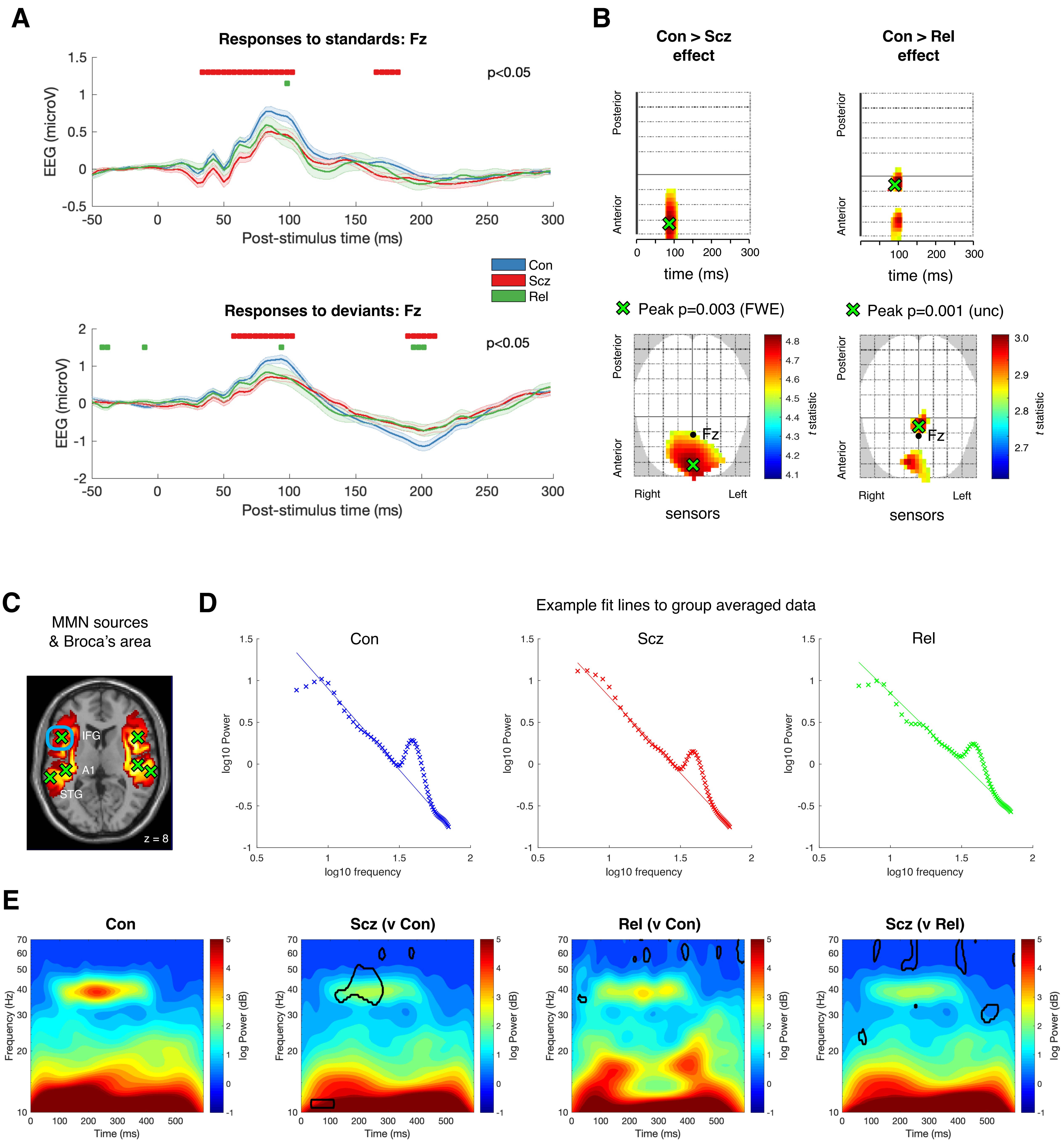

**Figure S2 - Additional MMN & 40 Hz ASSR (not normalized) results**

A – Standard (above) and deviant (below) MMN responses for Con (n=94), Scz (n=96) and Rel (n=42) at electrode Fz. Red bars denote group differences in Con vs Scz, green bars denote group differences in Con vs Rel, both $P<0.05$ ($t$-tests at each timepoint, uncorrected).

B – Left: The lower plot shows the overall Con > Scz effect at sensor level, displayed at $P<0.05$ (FWE). The significant cluster highlights the group differences common to both standard and deviant conditions at around 100 ms (also visible in panel A). The location of electrode Fz is shown in black. The peak effect is shown in green: $P=0.003$ (FWE), $t(376)=4.83$. The upper plot is of anterior to posterior sensors vs time: the peak effect occurs at 82 ms.

Right: These plots show the overall Con > Rel effects at sensor level in the same format, displayed at $P<0.005$ (unc); the peak effect is at 94 ms, $P=0.001$ (unc), $t(268)=3.02$.

C – Group source localization of the MMN data, showing the three bilateral sources used in the DCM model (green crosses). NB this source localization is just for illustrative purposes: source localization was performed for each subject as part of the DCM inversion. Broca's area (left BA44 and BA45) is highlighted in the blue box.

D – These plots show the mean 40 Hz ASSR power spectra at electrode Fz for Con (n=92), Scz (n=94) and Rel (n=42). Each plot shows an example line of best fit (using robust fitting to the 10-30 Hz and 50-70 Hz ranges). These were used on an individual-subject basis to normalize their spectra by subtracting this 1/f gradient.

E – Unnormalized time frequency plots for the 40 Hz ASSR paradigm; the three rightmost plots show areas of group differences at the $P<0.05$ level (encircled) at each time point and frequency. Without normalization, there is no significant difference between Con and Rel at 40 Hz, but with normalization there is (Figure 4C).

### Fig S3: MMN, 40 Hz ASSR & rsfMRI model fits

**A**

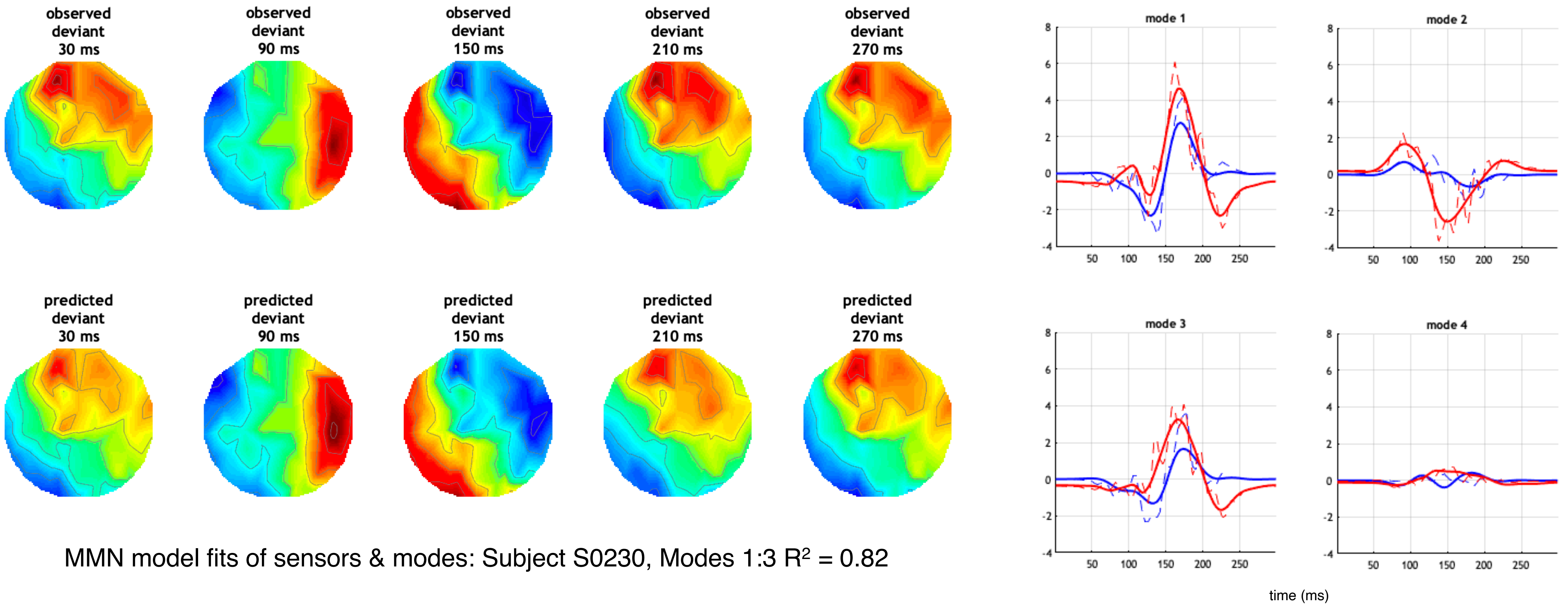

MMN model fits of sensors & modes: Subject S0230, Modes 1:3 $R^2 = 0.82$

**B**

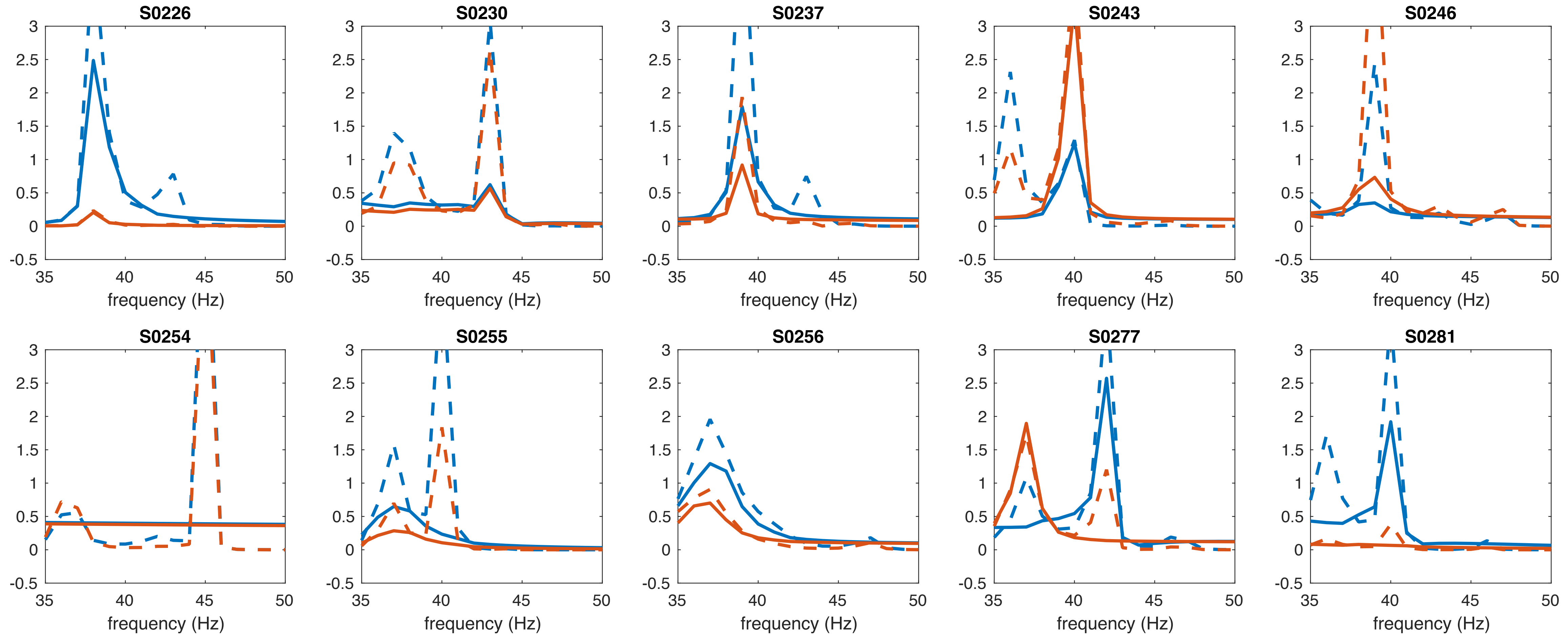

**C**

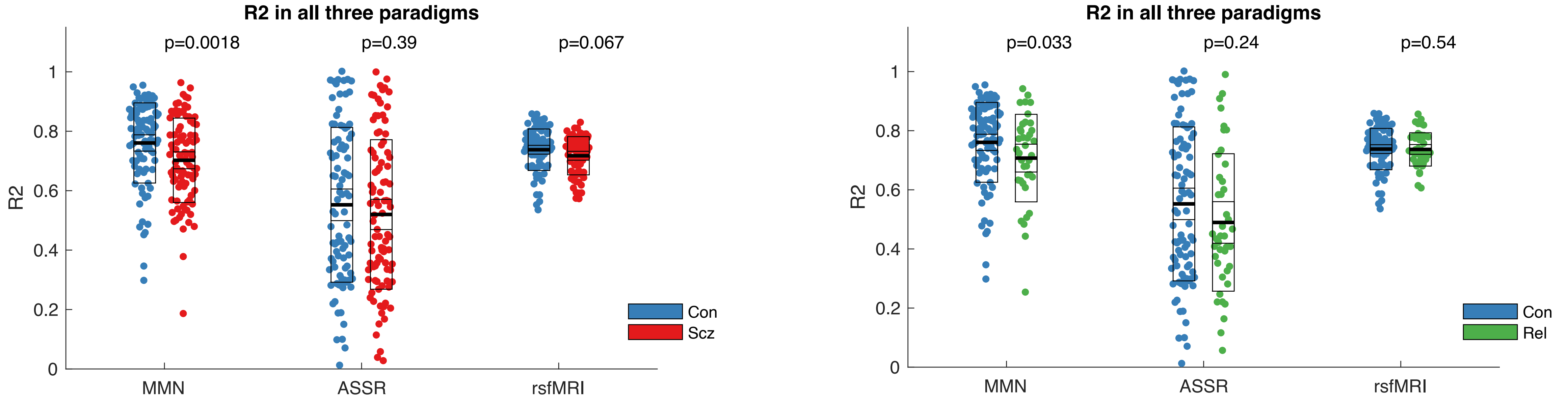

**Figure S3 – MMN, 40 Hz ASSR and rsfMRI DCM model fits**

A – Left: Example MMN model fits from a single subject (S0230) in sensor space. The top row shows the EEG data in the deviant condition at various timepoints; the bottom row shows the corresponding EEG data generated by the model parameters.

Right: Example MMN model fits of the first four (of eight) modes of the prior predicted covariance in sensor space. The standard and deviant conditions are plotted in blue and red respectively, data are plotted in dashed lines, the model fits in full lines. The first three modes capture most of the variance in the data.

B – 40 Hz ASSR model fits (full lines) for the first 10 subjects, for the power spectra (dashed lines) in left A1 (blue) and right A1 (orange). Fits are generally satisfactory – an exception is S0254, whose peak at >45 Hz is too far from the prior of 40 Hz for the model to accommodate.

C – These plots compare $R^2$ values for all three paradigms in Con and Scz (left) and Con and Rel (right) using ranksum tests as the distributions are skewed. *P* values are not corrected for multiple comparisons. In the MMN, Con have higher $R^2$ values than Scz ($Z$=3.16, $P$=0.0018), and Rel ($Z$=2.14, $P$=0.033). There are no significant differences in the other two paradigms.

### Fig S4: MMN DCM model spaces and additional results

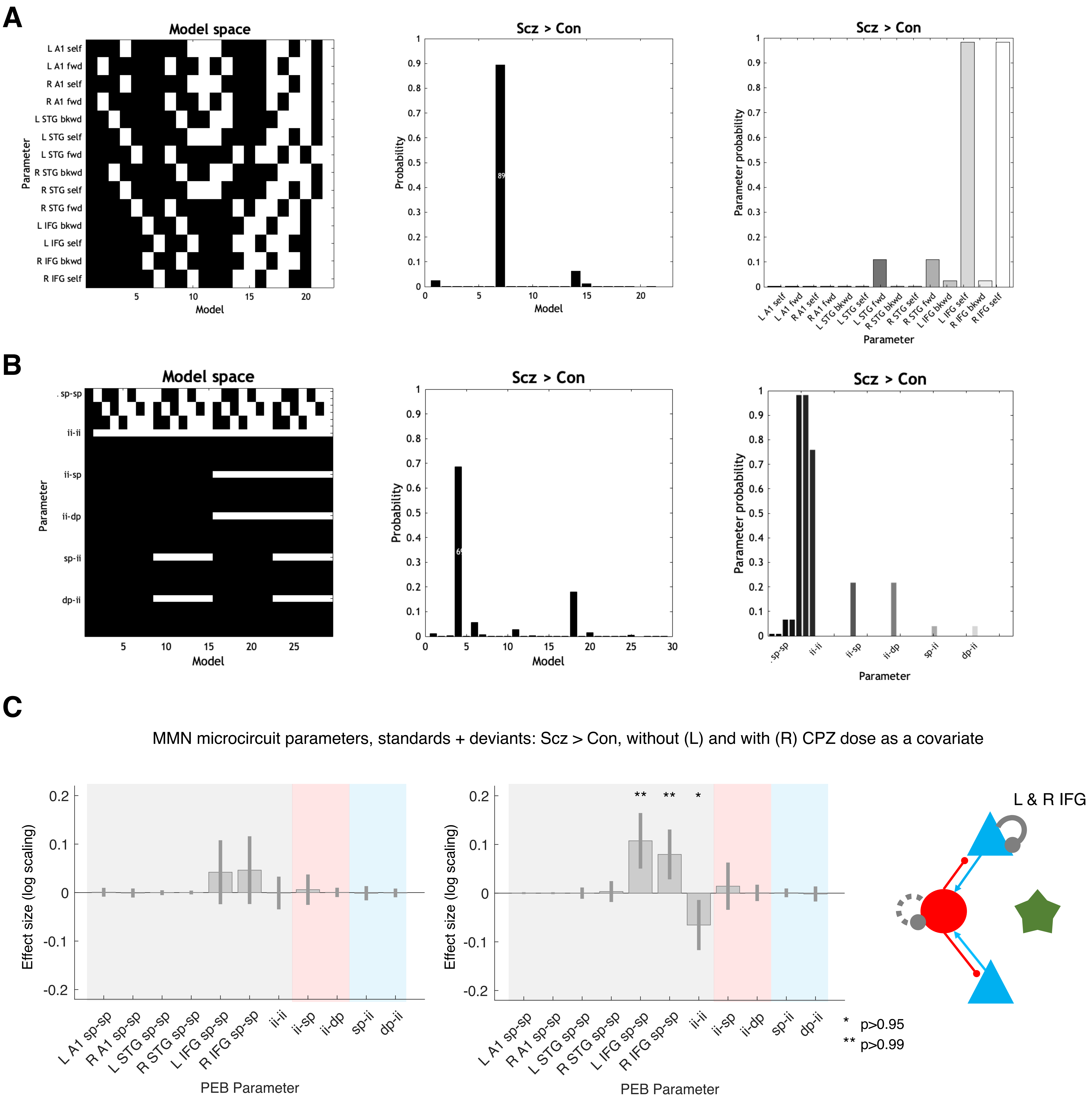

**Figure S4 – MMN DCM model spaces and additional results**

A – Left: The model space used to analyse the Scz > Con mismatch effect (and all other mismatch effects) within the PEB framework. The 22 rows each represent a model containing different combinations of extrinsic or self-inhibitory connection parameters (in white) that could best explain the group difference in mismatch effects. The first model is a null model, i.e. that no parameters differ between groups.

Middle: The posterior probabilities of each of the 22 models are shown, following Bayesian model reduction. Model 7 (containing only bilateral IFG self-inhibitory connections) is the best model with a posterior probability of $P$=0.89.

Right: The posterior parameter probabilities are shown, following Bayesian model averaging. These are the probabilities that each parameter contributes to the group difference effect. The only two parameters of $P$>0.95 are self-inhibition in bilateral IFG.

B – These plots show the PEB analysis of the overall Scz > Con effect in the MMN (i.e. across both conditions) in the same format as above. There are 11 parameters in the model space (the same was used for all MMN microcircuit analyses): six sp-sp self-inhibitory parameters (one per region) and five more microcircuit parameters (identical in every region, hence they each appear only once in the rows). The results pertain to the analysis shown in Figure S4C (right).

C – These plots show overall Scz > Con differences in microcircuit parameters in the MMN (i.e. across both standard and deviant conditions), without (left) and with (right) the inclusion of chlorpromazine dose equivalent (CPZ) as a covariate of no interest, in the same format as Figure 3F. Without CPZ, the IFG effects only reach the $P$>0.75 level. With the inclusion of CPZ, however, Scz show increased sp self-inhibition (i.e. reduced synaptic gain) in bilateral IFG, and disinhibition of interneurons. CPZ is thus unlikely to be contributing to the increased sp self-inhibition seen in Scz: if anything, it might be ameliorating this deficit. These L and R IFG effects are also present without the addition of age, sex and smoking covariates ($P$>0.99).

### Fig S5: 40 Hz ASSR DCM model space and additional results

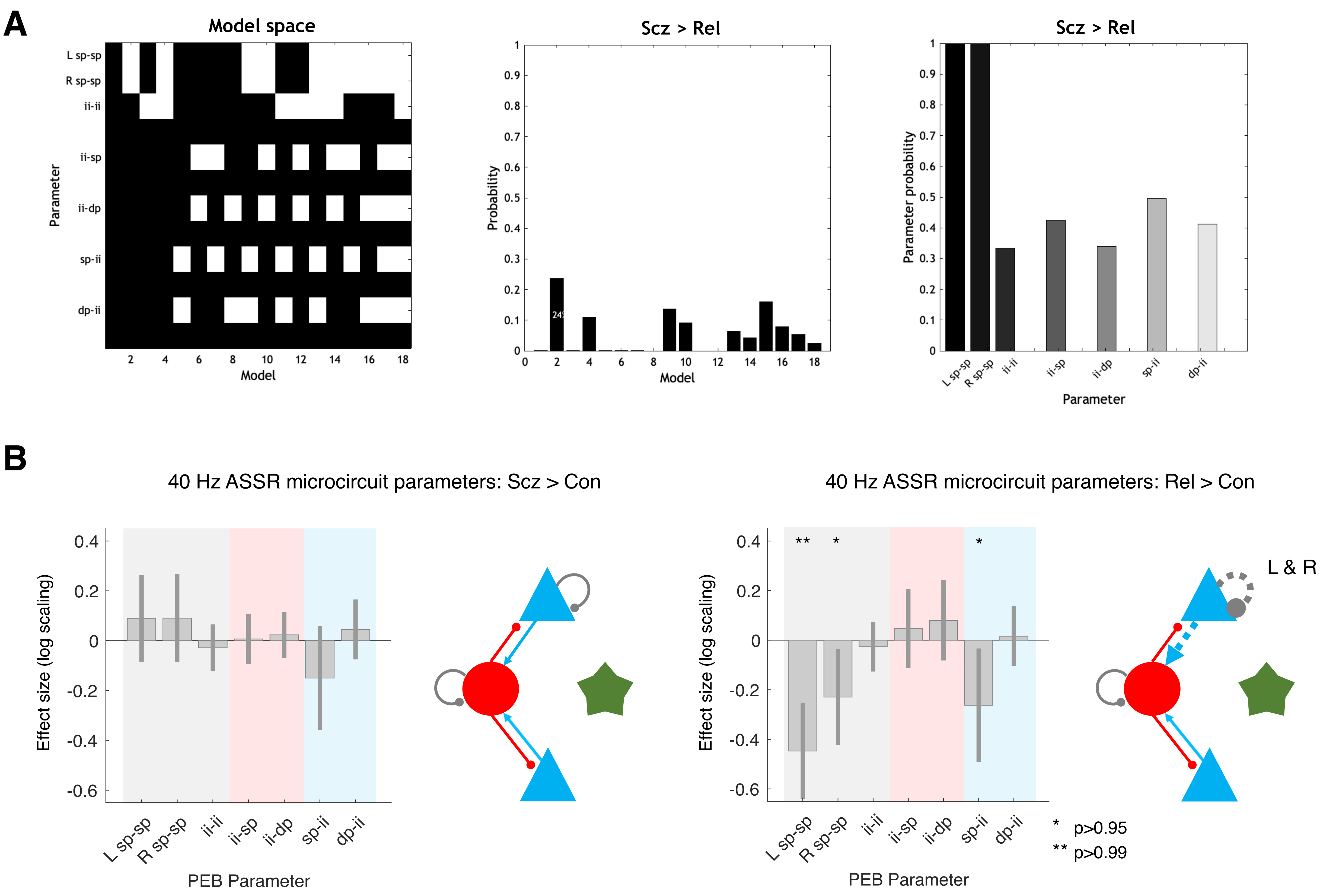

**Figure S5 – 40 Hz ASSR DCM model space and additional results**

A – These plots show the PEB analysis of the Scz > Rel effect in the 40 Hz ASSR in the same format as Figure S4A. The model space (identical in every 40 Hz ASSR analysis) contains different combinations of seven parameters: sp self-inhibition in L and R A1, and the five other microcircuit parameters (each constrained to be the same in both areas). No model is very likely (middle plot) but the L and R sp self-inhibition parameters appear in every one of the probable models, hence they are very likely differing between groups (right plot).

B – Left: The Scz > Con contrast did not show effects of $P>0.95$.

Right: Interestingly, the Rel > Con contrast indicated a loss of sp input into interneurons but also a disinhibition of sp cells – the former effect was also seen at $P>0.75$ in Scz, but the latter effect was reversed in Scz at $P>0.75$: see Figure 3C for an analysis combining all three groups. All effects shown are also present without the addition of age, sex and smoking covariates ($P>0.95$).

### Fig S6: rsfMRI DCM model space and additional results

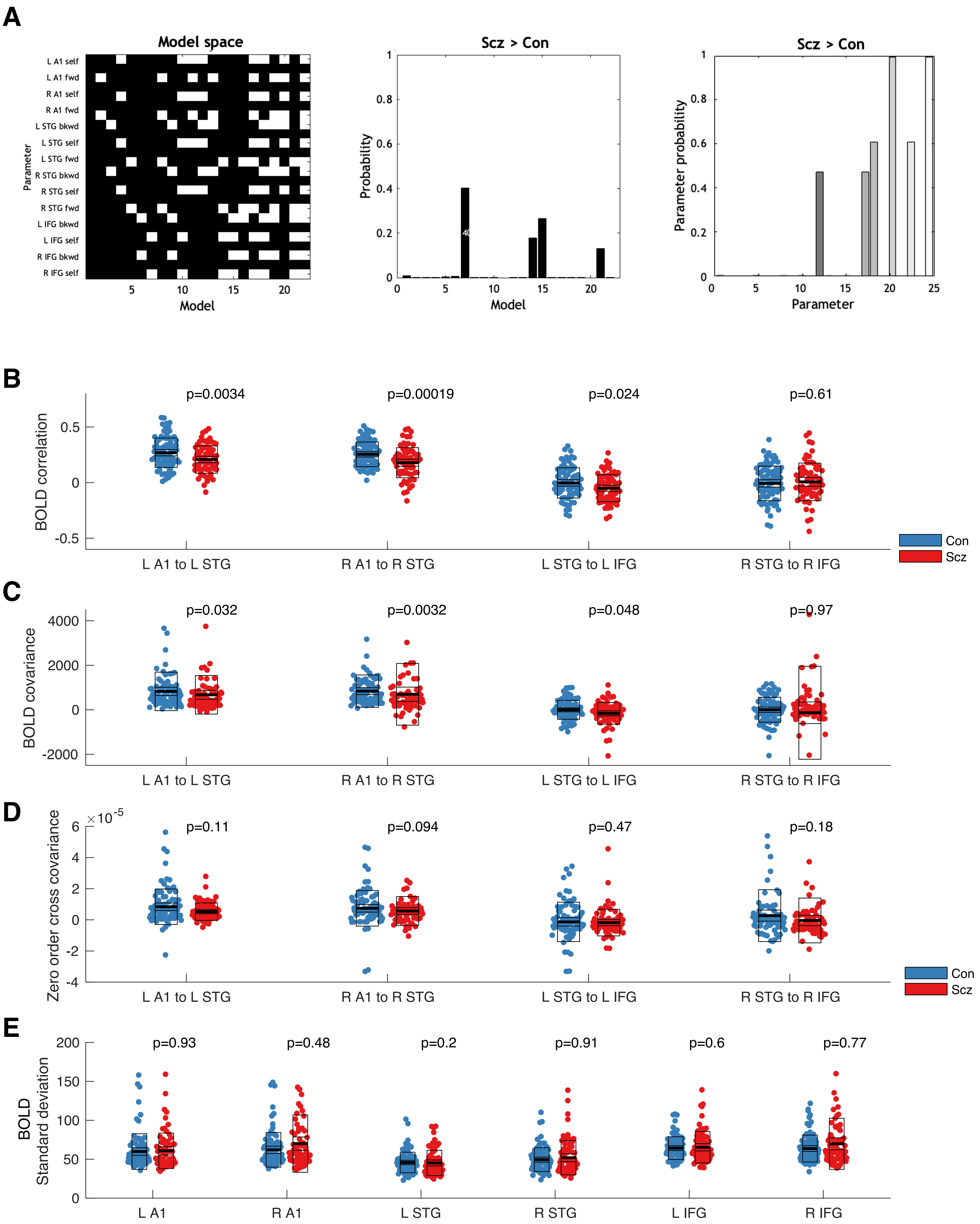

**Figure S6 – resting state fMRI DCM model space and additional results**

A – These plots show the PEB analysis of the Scz > Con effect in the rsfMRI paradigm in the same format as Figure S4A. The model space (identical in every rsfMRI analysis) contains different combinations of forward, backward and self-inhibitory connections in the same cortical network used in the MMN analysis.

B – Functional connectivity analysis (with global signal regression, GSR) between nodes of the MMN network in Scz (n=72) vs Con (n=85). There are significant decreases in functional connectivity (i.e. Pearson correlation) between A1 and STG bilaterally in Scz, and between L STG-IFG, although the latter would not survive multiple comparison correction (L A1-STG $t(154)=2.97$, $P=0.0034$; R A1-STG $t(154)=3.83$, $P=0.00019$; L STG-IFG $t(154)=2.29$, $P=0.024$, $P$ values not Bonferroni-corrected). Note that these findings are quite different from the results of effective connectivity analysis in Figure 5A.

C – The functional connectivity analysis (with GSR) is repeated using covariance instead of correlation, and ranksum tests as the distributions are skewed. Scz vs Con differences are less marked (L A1-STG $Z=2.15$, $P=0.032$; R A1-STG $Z=2.95$, $P=0.0032$; L STG-IFG $Z=1.96$, $P=0.048$, $P$ values not Bonferroni-corrected).

D – For comparison, the covariance (at zero lag, as in functional connectivity analysis) of the underlying neuronal activity inferred by DCM in each source is shown. There are no longer group differences (ranksum tests, all $P>0.05$).

E – Standard deviations of the BOLD timeseries (with GSR) in each of the nodes of the network – no nodes show group differences (ranksum tests, all $P>0.1$), including L and R IFG, where self-inhibition is greater in Scz (Figure 5A).

See the Online Methods for a discussion of how inferred effective connectivity may or may not be related to the corresponding functional connectivity between nodes (or variance within a node).

### Fig S7: rsfMRI results without GSR

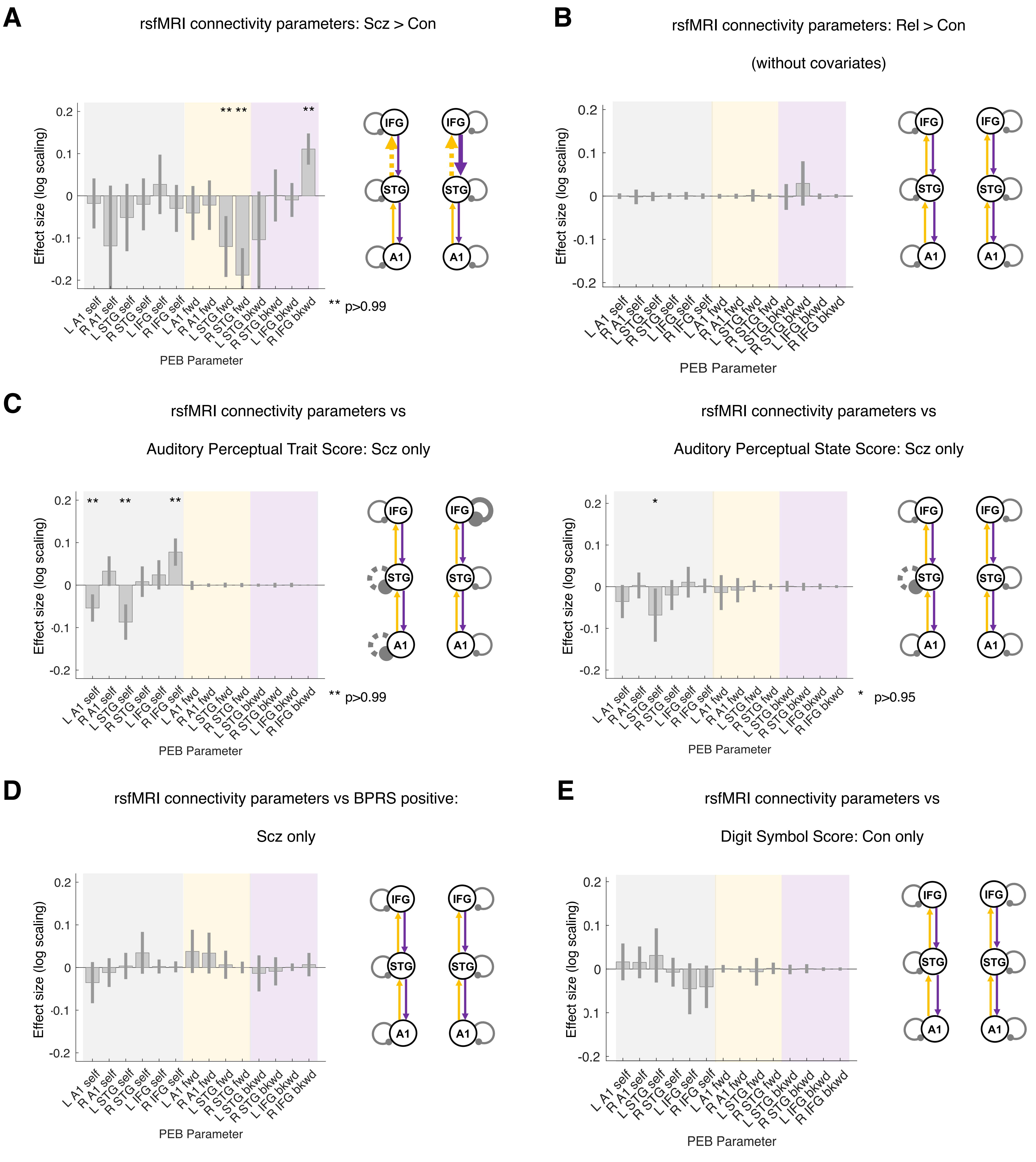

**Figure S7 – Resting state fMRI results without global signal regression**

These plots (in the same format as Figure 5) illustrate the importance of the global signal regression (GSR) preprocessing step in the rsfMRI analysis of Scz (n=72) and Con (n=85): many results change substantially without it.

A – With GSR, Scz show a loss of gain in bilateral IFG, but without GSR, Scz show a loss of forward frontotemporal connectivity instead.

B – With GSR, Rel show a loss of gain in bilateral IFG (without inclusion of the age covariate), but without GSR, no effects are significant (without the age covariate).

C – Left: With GSR, abnormal auditory percepts 'trait' score in Scz relate to loss of gain in bilateral IFG; without GSR, this effect is still seen in R IFG, but there are also effects of disinhibition in L temporal cortex (similar to state abnormal auditory perceptions both with and without GSR).

Right: With GSR, abnormal auditory percepts 'state' score in Scz relate to disinhibition in bilateral temporal lobes and loss of STG-A1 connectivity also; without GSR, only the disinhibition in L STG survives.

D – With GSR, BPRS positive symptoms related to disinhibition in five regions and increased forward connectivity in three connections; without GSR, only disinhibition in L A1 and increased forward connectivity from bilateral A1 to STG are seen, at $P>0.75$.

E – With GSR, Digit Symbol performance in the Con group relates to R IFG excitability; this is also the case without GSR, in bilateral IFG, but only at $P>0.75$.

### Fig S8: EEG parameter sensitivity analysis

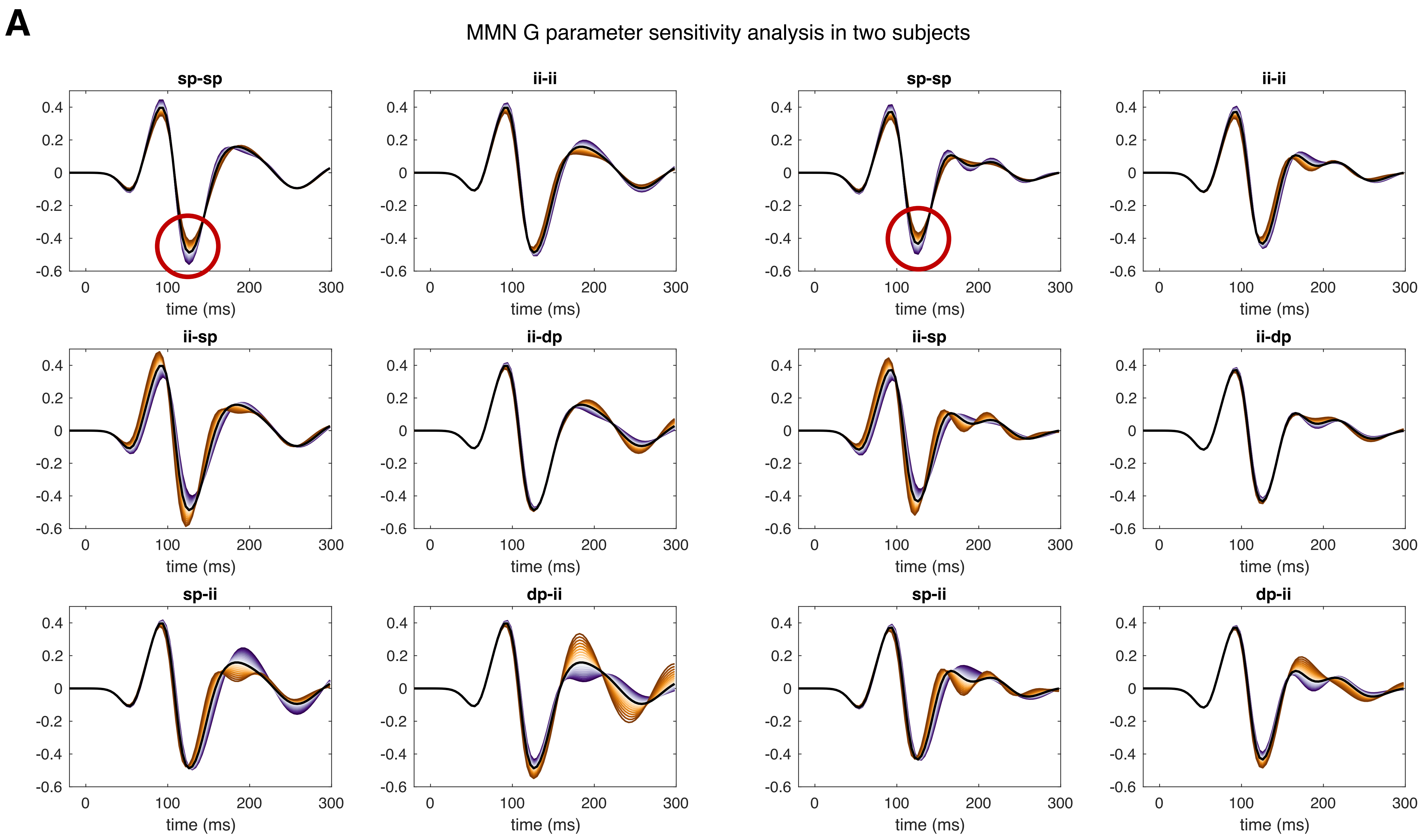

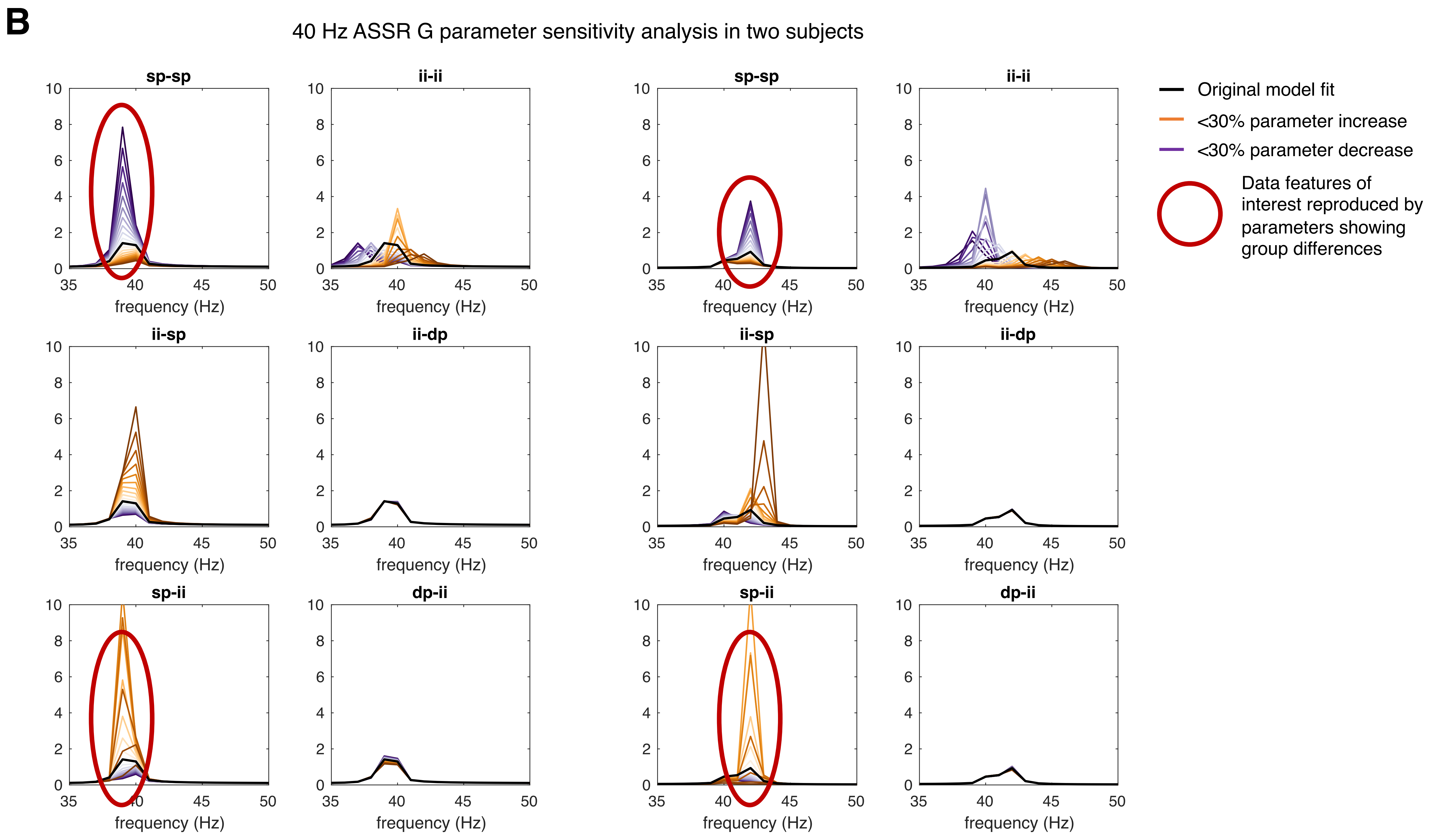

**Figure S8 – EEG microcircuit parameter sensitivity analyses**

This figure shows the results of simulating virtual electrode data in a single cortical area to reproduce key data features in the MMN and 40 Hz ASSR paradigms. In each paradigm, two subjects representative of the group average were selected, and their posterior G (intrinsic connectivity) parameters used to simulate data. In each simulation, one G parameter in turn was progressively changed by ±30% – results of increases and decreases are shown in orange and purple, with darker shades representing bigger changes. The left and right panels correspond to the two subjects in each paradigm, and the six plots in each panel to the six G parameters: sp-sp, ii-ii, ii-sp, ii-dp, sp-ii and dp-ii connectivities. Note that the first two are self-inhibitory connections (synaptic gain).

A – Sensitivity analysis in the MMN paradigm. The ERP data showed a reduced MMN amplitude (Figure 3A) and P100 amplitude (Figure S2A) but no significant change in MMN latency in Scz; in Scz, sp-sp was increased (Figure 3F). In both subjects' simulations, increasing sp-sp (i.e. reducing synaptic gain) reduces the amplitude of the negative deflection (circled in red) without changing the latency, as it also does to the P100. Increasing ii-ii has similar but smaller effects on these amplitudes, but shortens the latency. Decreasing ii-sp likewise reduces the amplitudes, but lengthens the latency. The remaining parameters cause more substantial changes in the latter half of the waveform. The waveform itself is compressed in time compared with empirical data, because extrinsic connections – which cause a ~10 ms delay per connection – are not included in the simulation of a single area.

B – Sensitivity analysis in the 40 Hz ASSR paradigm. The power spectral data showed a reduction in ~40 Hz power (Figure 4C). In both subjects' simulations, increasing sp-sp and decreasing sp-ii (the parameter changes in Scz vs Rel and Scz+Rel vs Con respectively – Figure 4G) both reduce ~40 Hz power (circled in red). Reducing ii-sp has similar but less powerful effects on sp-ii. Changing ii-ii changes the peak frequency, but its effects on power are only prominent at precisely 40 Hz. Dp-ii and ii-dp connections have minimal effects.

### Fig S9: Scz > Con differences by age group

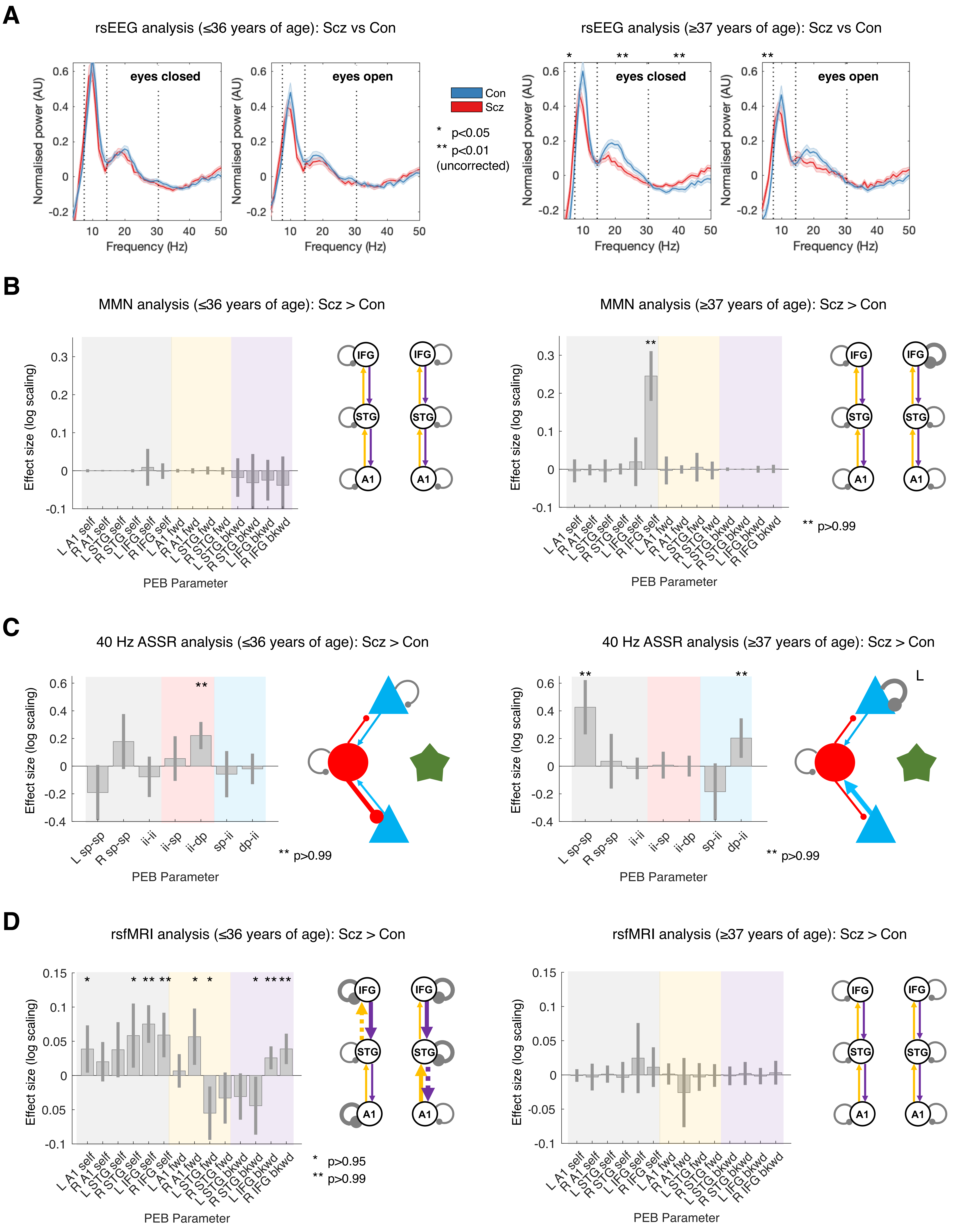

**Figure S9 – Scz > Con group differences in younger and older subjects**

This figure illustrates the key Scz > Con group difference analyses from the four paradigms, with both groups divided according to median age. The aim was to investigate whether Scz with more recent or more longstanding diagnoses were driving the group difference effects detected previously.

A – rsEEG power spectra in eyes open and closed conditions, plotted in the same format as Figure 2A. Group differences in total power in each frequency band are indicated with asterisks (ranksum tests, not corrected for multiple comparisons). Left: rsEEG in subjects ≤36 years old (Con n=50, Scz n=49), there were no detected differences in any frequency band in either eyes open or closed conditions (ranksum tests, all $P$>0.05, uncorrected for multiple comparisons). Right: rsEEG in subjects ≥37 years old (Con n=48, Scz n=46), in the eyes closed condition, Scz had higher θ ($Z$=2.24, $P$=0.025), lower β ($Z$=-2.92, $P$=0.0035) and higher γ ($Z$=2.58, $P$=0.0098), but no difference in α power ($Z$=-1.35, $P$=0.18). In the eyes open condition, Scz had higher θ ($Z$=3.27, $P$=0.001), but no significant differences in α ($Z$=-0.9, $P$=0.4), β ($Z$=-1.78, $P$=0.075) or γ power ($Z$=1.93, $P$=0.053); all ranksum tests, uncorrected for multiple comparisons. Older Scz are driving the effects in Figure 2B.

B – PEB analyses of the MMN mismatch contrast, plotted in the same format as Figure 3F. Left: In subjects ≤36 years old (Con n=45, Scz n=47), no parameters differed between groups (all $P$<0.95). Right: In subjects ≥37 years old (Con n=48, Scz n=48), R IFG self-inhibition was higher in Scz ($P$>0.99). Older Scz are driving the R IFG effect in Figure 3F.

C – PEB analyses of the 40 Hz ASSR, plotted in the same format as Figure 4G. N.B. there were no group differences in the original Scz > Con contrast (Figure S5B), but there were in the Scz > Rel (psychosis 'state') effect (Figure 4G). Left: In subjects ≤36 years old (Con n=47, Scz n=48), ii-dp connectivity was increased in Scz ($P$>0.99). In subjects ≥37 years old (Con n=46, Scz n=46), dp-ii and L A1 self-inhibition were increased in Scz (both $P$>0.99). Older Scz are driving the L A1 self-inhibition effect in Figure 4G.

D – PEB analyses of the rsfMRI, plotted in the same format as Figure 5A. Left: In subjects ≤36 years old (Con n=44, Scz n=34), numerous nodes show increases in self-inhibition in Scz, including bilateral IFG (both $P$>0.99), as well as changes in forward and backward connectivity. In subjects ≥37 years old (Con n=42, Scz n=37), there are no differences in parameters (all $P$<0.95). Younger Scz are driving the bilateral IFG self-inhibition effects in Figure 5A.

### Fig S10: rsfMRI & EEG DCM parameter relationships

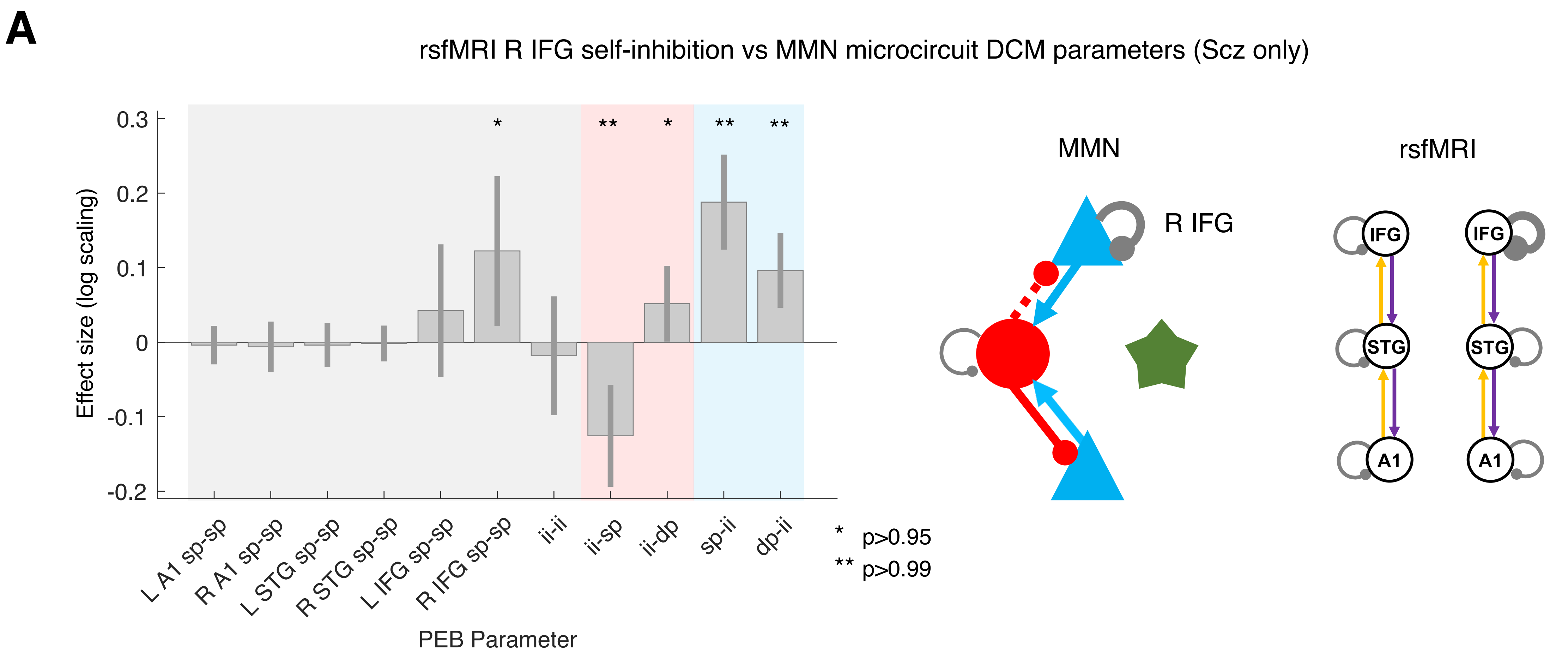

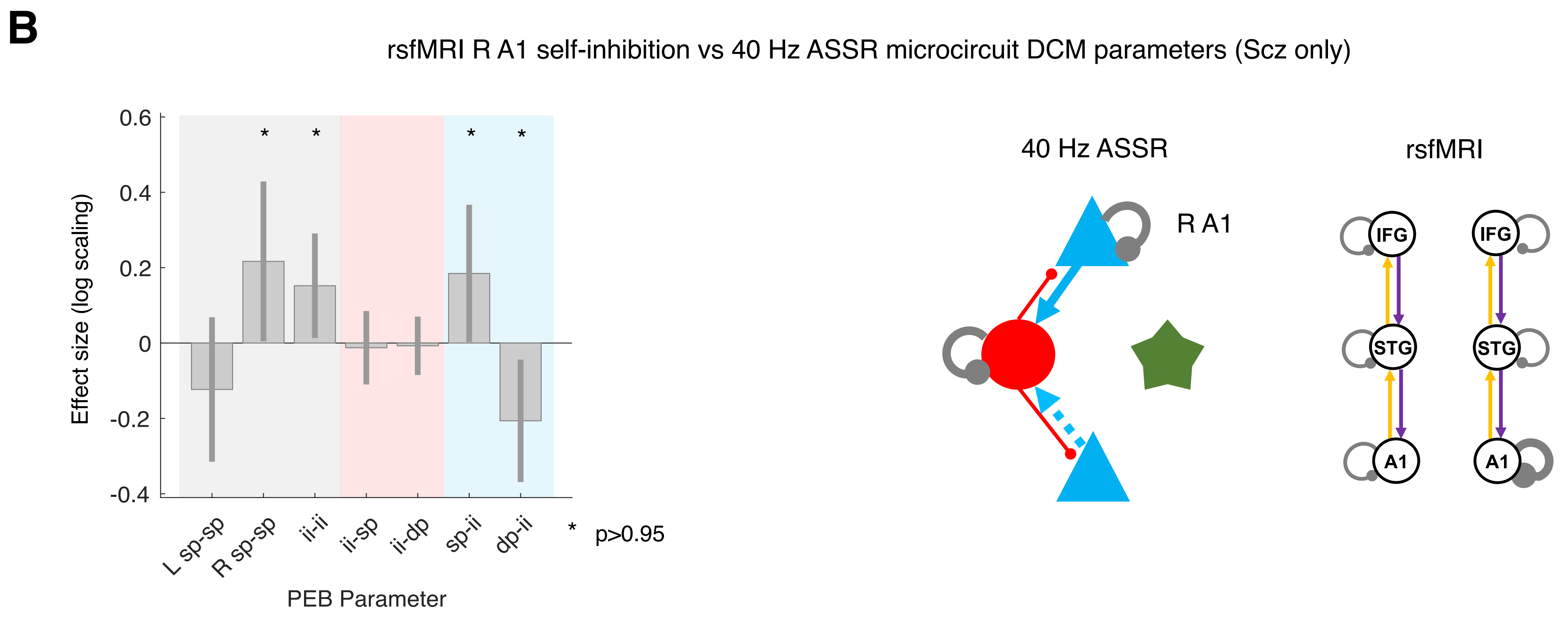

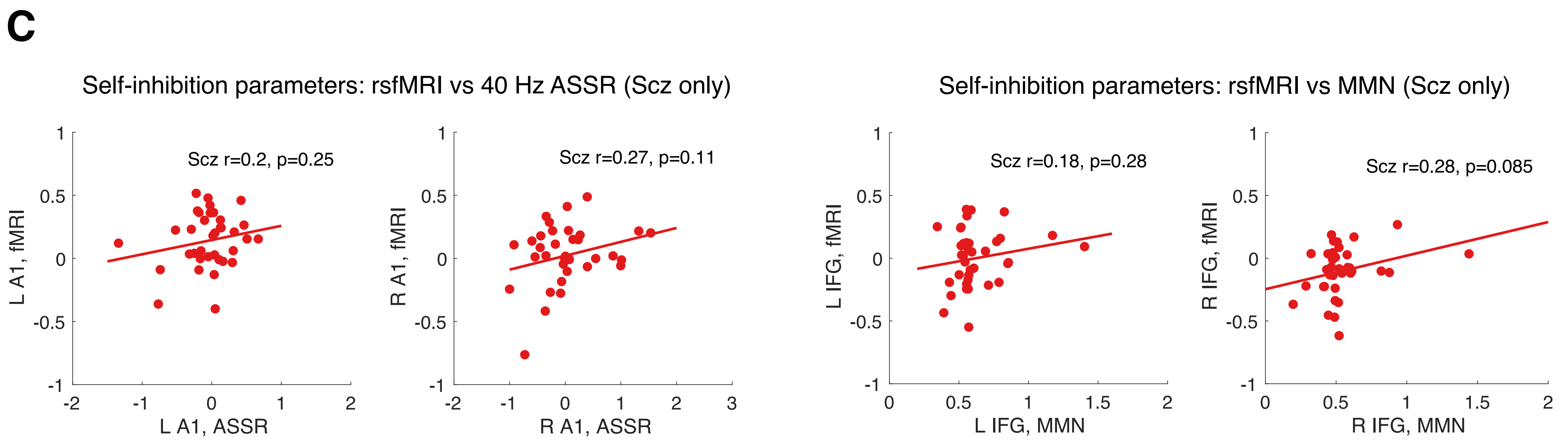

**Figure S10 – Resting state fMRI & EEG DCM parameter relationships in Scz**

All analyses are conducted on Scz whose rsfMRI and EEG paradigms were recorded <100 days apart (for MMN, n=44; for 40 Hz ASSR, n=40).

A – In Scz, increased self-inhibition in R IFG (estimated from rsfMRI) relates to both loss of superficial pyramidal cell gain in R IFG specifically and a net increase of inhibition of pyramidal cells (i.e. stronger pyramidal inputs to interneurons and stronger returning connections except for ii-sp, whose decrease is smaller than the sp-ii increase) in the MMN analysis. All effects (except the ii-dp increase) shown are also present without the addition of age, sex and smoking covariates ($P$>0.95).

B – In Scz, increased self-inhibition in R A1 (estimated from rsfMRI) relates to increased sp self-inhibition in R A1 specifically and stronger sp input to interneurons. It also – counterintuitively – relates to greater self-inhibition of and weaker dp input to interneurons. All effects (except the dp-ii decrease) shown are also present without the addition of age, sex and smoking covariates ($P$>0.95).

NB With the inclusion of chlorpromazine dose equivalents as a covariate the specific R IFG effect (in A) and R A1 effect (in B) are lost, however, and likewise if Scz with rsfMRI and EEG paradigms >100 days apart are included.

C – These plots show the correlations between self-inhibition parameters across EEG and rsfMRI paradigms within Scz. The left two plots show correlations between 40 Hz ASSR and rsfMRI estimates of self-inhibition in bilateral A1, the right two plots show correlations between MMN and rsfMRI estimates of self-inhibition in bilateral IFG. Relationships in R A1 and R IFG approached statistical significance, and are analysed in more detail using PEB in panels A and B above. There were no correlations between the variance of these parameters across these combinations of paradigms (all $P$>0.3).

#### Table S1: Participant characteristics

| | Controls | Scz | Relatives | Con v Scz | Con v Rel | Rel v Scz |
|---|---|---|---|---|---|---|
| Total | 107 | 108 | 57 | | | |
| **Demographics** | | | | | | |
| Age, years (mean ±std) | 39.4 ±14.3 | 39.4 ±13.9 | 45.4 ±16.6 | p=1, t=0.0 | **p=0.02, t=2.4** | **p=0.01, t=2.5** |
| Sex | 64 M, 43 F | 73 M, 35 F | 16 M, 41 F | p=0.2, $\chi$=1.4 | **p=$10^{-4}$, $\chi$=15** | **p=$10^{-6}$, $\chi$=23** |
| Smoking (current) | 35 (33%) | 42 (39%) | 9 (16%) | p=0.3, $\chi$=0.9 | **p=0.02, $\chi$=5.4** | **p=$10^{-3}$, $\chi$=9.3** |
| **Psychological measures** | | | | | | |
| Digit Symbol (mean ±std) | 10.5 ±3.1 | 7.6 ±3.0 | 10.2 ±3.1 | **p=$10^{-9}$, t=6.5** | p=0.6, t=0.5 | **p=$10^{-6}$, t=4.9** |
| APTS Trait (mean ±std) (min 0, max 48) | 4.4 ±5.4 | 18.2 ±13.4 | 4.2 ±4.6 | **p=0, t=9.8** | p=0.8, t=0.2 | **p=$10^{-11}$, t=7.5** |
| APTS State (mean ±std) (min 0, max 48) | 0.9 ±2.0 | 10.0 ±11.7 | 1.5 ±3.3 | **p=$10^{-13}$, t=7.9** | p=0.2, t=1.2 | **p=$10^{-7}$, t=5.3** |
| **Clinical measures** | | | | | | |
| Antipsychotic prescribed | 0 | 90 | 1 | | | |
| Antipsychotic types | 67 atypical*, 15 typical*, 22 Clozapine*, 3 unmedicated, 15 not recorded<br>*includes 15 on combinations of these | | | | | |
| CPZ equivalent, mg (mean ±std) | n/a | 464 ±488 | 3.5 ±26.3 | | | |
| Antidepressant prescribed | 13 | 39 | 9 | | | |
| Benzodiazepine (not daily) prescribed | 5 | 16 | 3 | | | |
| Mood stabiliser prescribed | 3 | 21 | 1 | | | |
| BPRS total (mean ±std) (scale from 20 to 140) | n/a | 38.8 ±12.3 | n/a | | | |
| BPRS positive (mean ±std) (scale from 7 to 49) | n/a | 14.4 ±7.4 | n/a | | | |
| BPRS negative (mean ±std) (scale from 4 to 28) | n/a | 7.3 ±4.5 | n/a | | | |
| **Imaging paradigms** | | | | | | |
| rsEEG analysed/obtained | 98/107 | 95/108 | n/a | p=0.8, $\chi$=0.4 | | |
| MMN modelled/obtained | 93/98 | 95/100 | 40/48 | p=1, $\chi$=0.0 | **p=0.02, $\chi$=5.3** | **p=0.02, $\chi$=5.3** |
| ASSR modelled/obtained | 92/101 | 94/105 | 42/53 | p=0.7, $\chi$=0.1 | **p=0.04, $\chi$=4.3** | p=0.08, $\chi$=3.1 |
| rsfMRI modelled/obtained | 85/107 | 72/108 | 45/54 | **p=0.03, $\chi$=4.5** | p=0.6, $\chi$=0.4 | **p=0.03, $\chi$=5.0** |
| rsfMRI motion scrubbed frames, % (mean ±std) | 4.1 ±6.2 | 6.4 ±9.9 | 5.0 ±8.6 | p=0.08, t=1.7 | p=0.5, t=0.7 | p=0.5, t=0.7 |
| rsfMRI SNR | 46.6 ±9.2 | 43.5 ±13.0 | 43.9 ±10.7 | p=0.09, t=1.7 | p=0.1, t=1.5 | p=0.9, t=0.2 |
| Days between rsfMRI & MMN | 72 ±217 | 64 ±222 | 27 ±140 | | | |
| <100 days between rsfMRI & MMN modelled/obtained | 55/74 | 44/73 | 28/40 | | | |
| Days between rsfMRI & ASSR | 67 ±162 | 106 ±258 | 35 ±158 | | | |
| <100 days between rsfMRI & ASSR modelled/obtained | 60/79 | 40/81 | 35/45 | | | |
